# Risk Factors for Severe Post-COVID Condition in Children, Adolescents, and Young Adults

**DOI:** 10.64898/2025.12.01.25341171

**Authors:** Quirin Donath, Matthias Haegele, Daniela Schindler, Tiziana Welzhofer, Catharina Christa, Annika Grabbe, Ariane Leone, Clara Ilhan, Carola Weidmann, MA Maria Eberhartinger, Sara Bechtold, Nicola Bursch, Hedwig Wolf, Hannah Hieber, Laura-Carlotta Peo, Lara A. Bucka, Silvia Stojanov, Cordula Warlitz, Martin Alberer, Katrin Gerrer, Anna Hausruckinger, Kirstin Mittelstrass, Clemens-Martin Wendtner, Manuela A. Hoechstetter, Armin Grübl, Nicole Toepfner, Rafael Pricoco, Carmen Scheibenbogen, Lorenz L. Mihatsch, Uta Behrends

**Author notes:** **Corresponding author:** Lorenz Mihatsch MD, MSc, TUM University Hospital of the Technical University of Munich Ismaninger Str. 22, 81675 Munich Germany. These authors contributed equally to this work (Lorenz L. Mihatsch, Uta Behrends).

## Abstract

**Background:** Post-COVID condition (PCC) in children and young people (PCCcyp) remains a significant health burden. Early identification of patients at risk for severe disease, including myalgic encephalomyelitis/ chronic fatigue syndrome (ME/CFS), is crucial to provide timely and adequate care.

**Methods:** This monocentric, observational registry study was performed at a tertiary pediatric hospital in Germany. Children and young people aged 7–25 years with post-COVID condition (PCC), according to the WHO, were included at the time of diagnosis. Standardized clinical assessment tools and patient-reported outcome measures were applied, including the novel Munich Long COVID Symptom Questionnaire (MLCSQ). Severe PCC was defined by clustered chronic symptom burden, fatigue severity scale (FSS), Total Composite Autonomic Symptom Score-31 (COMPASS-31), SF-36 composite scores, Bell Score, and confirmed ME/CFS diagnosis.

**Findings:** Of 120 participants, severe PCCcyp was associated with a higher number of acute symptoms (adjusted OR 1·22), acute orthostatic intolerance (adjusted OR 9·87), acute trouble concentrating (adjusted OR 11·8), and female sex (OR 3·31). Categorising acute symptoms at a threshold of ≥12 yielded the best model performance (AUC 0·857; sensitivity 65·6%; specificity 90·2%). ME/CFS was diagnosed in 24% participants, all within the severe PCC cluster, and was characterised by greater acute symptom complexity, more fatigue, more autonomic symptoms, and poorer physical function.

**Interpretation:** The number of acute symptoms and/or individual symptoms during the acute phase of the SARS-CoV-2 infection serve as early and specific predictors for severe PCCcyp. In particular, patients with ≥12 acute symptoms should be closely monitored to enable early diagnosis of severe PCC and potentially ME/CFS. A distinct cluster of severely affected patients – frequently with ME/CFS diagnosis – was identified.

**Funding:** This study was funded by the Bavarian State Ministry of Health and Care, and the German Center for Infection Research.

**Research in context Evidence before this study:** Post-COVID condition (PCC) in children and young people (CYP) has been increasingly recognized, but robust evidence on predictors of severe disease has remained scarce. Previous reports in adult patients documented a wide spectrum of persistent symptoms, frequently including fatigue, neurocognitive complaints, and autonomic dysfunction, yet no study systematically investigated acute-phase predictors for severe PCCcyp.

**Added value of this study:** In this monocentric observational registry study on 120 CYP aged 7–25 years with PCC, we identified acute-phase risk factors for severe pediatric PCC. We show that a high number of acute symptoms (≥12 symptoms), female sex, acute orthostatic intolerance, and/or acute trouble concentrating strongly predicted a severe PCCcyp cluster. Importantly, 24% of patients fulfilled ME/CFS diagnostic criteria, and all clustered within the severe PCC group, underscoring the clinical overlap between severe pediatric PCC and ME/CFS.

**Implications of all the available evidence:** These findings provide a framework for early risk stratification in pediatric PCC. Recognising acute predictors is crucial to provide timely and adequate care, particularly to those with 12 or more acute symptoms and/or specific symptoms during the acute SARS-CoV-2 infection. The identification of a clinically relevant ME/CFS subgroup underscores the need to integrate ME/CFS expertise into pediatric PCC care pathways. Together, this evidence can inform clinical management, healthcare planning, and the design of interventional trials targeting the most vulnerable CYP with PCC.

## INTRODUCTION

Post-acute sequelae of coronavirus disease 2019 (COVID-19) (PASC), often referred to as Long COVID, represent major clinical and economic challenges worldwide.^1–4^ They include ongoing, relapsing, or new symptoms at four weeks or later after infection with severe acute respiratory syndrome coronavirus 2 (SARS-CoV-2).^5^ A chronic form of PASC has been defined by the WHO as Post-COVID Condition (PCC). According to the definition for children and adolescents, PCC occurs in individuals with a history of confirmed or probable SARS-CoV-2 infection, when experiencing symptoms lasting at least 2 months which initially occurred within 3 months of acute COVID-19.

PCC can cause symptoms across all organ systems and often significantly impacts functioning, participation, and health-related quality of life (HRQoL).^6^ Frequent symptoms include fatigue, exertional intolerance, headache, respiratory symptoms, cognitive dysfunction, including brain fog, sleep problems, loss of smell and taste, anxiety, and autonomic dysfunction.^7,8^ The latter can manifest as orthostatic intolerance (OI) with or without postural orthostatic tachycardia syndrome (PoTS).^9^ Some patients develop post-exertional malaise (PEM), a compromising condition with symptom worsening after daily activities that were tolerated before. Long-lasting PEM is a cardinal symptom of myalgic encephalomyelitis/chronic fatigue syndrome (ME/CFS), a chronic multisystemic disease defined by clinical criteria and mostly triggered by viral infections.^5,9,10^ Most recent studies estimate ME/CFS prevalences ranging from 4·5 to 11·6% in adult PCC patients, depending on the study design.^11^ Diagnosing PCC, PoTS, and ME/CFS requires a thorough diagnostic work-up to define clinical phenotypes and rule out other medical conditions with symptom overlap.

Data on PCC in CYP (PCCcyp) is still scarce, and age-specific factors are insufficiently understood.^5^ Depending on the study design, the reported PASC prevalence in minors ranged from 0·8% to 60%,^12^ with lower rates observed in studies with SARS-CoV-2-negative control groups.^6,12–15^ While most CYP recover within a few months, a subset develops long-term disease.^3,7,15,16^

Established risk factors for PCC in adults include infection with pre-Omicron variants,^3^ older age, female sex, pre-existing chronic conditions, lack of COVID-19 vaccination,^17^ more severe acute illness, and greater symptom complexity.^18^ Preliminary evidence suggests similar risk patterns in CYP,^19–22^ but risk factors of PCCcyp are more difficult to assess due to limited data and heterogenous age groups.

The IMMUC study on EBV-associated infectious mononucleosis (EBV-IM) and its long-term sequelae in CYP indicated that susceptibility to infection, a higher number of acute symptoms, and early gastrointestinal (GI) involvement predicted prolonged recovery and chronic fatigue.^23^ Especially, gastrointestinal symptoms even prior to EBV primary infection has been identified as a risk factor for severe ME/CFS.

Here, we report on clinical phenotypes during the acute and chronic phases after SARS-COV-2 infection of 120 CYP, including patients with post-COVID ME/CFS. We provide the first evidence for clinical risk factors of severe PCCcyp, which might facilitate timely identification and adequate care.

## METHODS

### Study Cohort and Case Definition

This monocenter observational registry study comprises medical data from 120 CYP aged 7 to 25 years and diagnosed with PCC according to WHO definitions^24^ between January 2021 and May 2024 at the Munich Chronic Fatigue Center for Young People (MCFC).

Confirmed SARS-CoV-2 infection was defined by positive PCR or rapid antigen test combined with typical COVID-19 symptoms, while probable SARS-CoV-2 infection was defined by COVID-19 symptoms after contact with a SARS-CoV-2 PCR-positive person and positive SARS-CoV-2 serology not explained by vaccination.

Confirmed MECFS was defined by fulfilling the criteria of the Institute of Medicine (IOM) and/or the Canadian Consensus Criteria (CCC) as assessed by the Munich Berlin Symptom Questionnaire (MBSQ)^9^ in a structured medical interview after symptom-oriented differential diagnosis. In case of pediatric “pediatric case definition” of L.A. Jason and colleagues (PCD-J) and/or the clinical diagnostic worksheet designed by P.C. Rowe and colleagues (CDW-R)^10^ were used.

### Assessed Data

Patients were asked to fill in questionnaires pre-admission, retrospectively addressing their and their family’s general medical history, their symptoms during the acute phase of the SARS-CoV-2 infection as well as their current symptoms, medical treatment, grade of participation in education/work, and HRQoL. To assess PCC symptoms, diagnostic ME/CFS criteria, severity of fatigue, presence, severity, and duration of PEM, autonomic symptoms, the daily activity level, and HRQoL we used the novel Munich Long COVID Symptom Questionnaire (MLCSQ), MBSQ^9^, the fatigue severity scale (FSS),^25^ the DePaul Symptom Questionnaire for Post-exertional Malaise (DSQ-PEM),^26^ the Composite Autonomic Symptom Score 31 (COMPASS 31),^27^ the Bell CFIDS disability scale (Bell Score), and the Short Form-36 Health Survey (SF-36) with Physical (PCS) and Mental Component Summary (MCS),^28^ respectively.

The MLCSQ was developed for structured age-independent assessment of PCC symptoms in 13 domains, encompassing 83 items (**Supplementary Material M1, Appendix pp.7-9**). Items were adapted from the WHO Global COVID-19 Clinical Platform Case Report Form (CRF) for PCC (2021), the Long COVID-19 Survey by the German Society for Pediatric Infectious Diseases (DGPI, 2021), and the COMPASS 31 (Sletten et al., 2012).^27^ Frequency and severity were quantitatively assessed using a Likert scale as “not anymore/intermittent/persistent” or “mild/moderate/severe”, respectively. Only symptoms rated as “intermittent” or “persistent” in frequency were considered as PCC symptoms in this study.

### Statistical Analysis

#### Descriptive Analysis

Continuous variables were described as median (interquartile range (IQR)) and compared between groups using Kruskal-Wallis or Wilcoxon Rank Sum tests. Categorical variables were described as absolute and relative frequencies (%) and tested with Chi-Squared or Fisher’s Exact tests as appropriate. A P-value < .05 was considered statistically significant.

#### Risk Factor Analysis Using Distinct Clinical Outcomes

Multivariable linear regression models for outcomes SF36-PCS, SF36-MCS, and the number of PCC symptoms, as well as logistic regression models for PEM, ME/CFS, and PoTS diagnosis, adjusted for sex and age, were used. Candidate predictor variables were COVID-19 vaccination status prior to infection,^17^ hospitalization during COVID-19,^29^ the number of acute symptoms,^23^ the presence of acute gastrointestinal^30^ or orthostatic intolerance symptoms, fatigue, and/or troubles concentrating during the acute SARS-CoV-2 infection.

#### Identification and Risk Factor Analysis for Severe PCC Cluster

A data-driven clustering approach was applied to identify patient subgroups based on PCC severity. The following outcome variables were used for the cluster definition: Bell Score, FSS, COMPASS 31 total score, SF36-PCS, SF36-MCS, number of PCC symptoms, ME/CFS diagnosis. A k-means clustering algorithm (k=2) was applied to the standardized data set. Two clusters encompassed patients with “moderate” and “severe” manifestations of these outcome variables, referred to as “moderate PCC” or “severe PCC”, respectively, for easy interpretability. The cluster membership was used as the primary outcome for subsequent analyses.

Using LASSO-penalized logistic regression, we assessed the relationship between features of the acute COVID-19 phase and subsequent cluster membership. Candidate predictor variables were the same as for the risk factor analysis using distinct clinical outcomes (see above).

The logistic LASSO regression model was fitted with 10-fold cross-validation to identify the optimal penalty parameter (λ) with the least deviance. To account for class imbalance between the “mild PCC” and “severe PCC” clusters, inverse class weights were applied.

The resulting coefficients of the cross-validated model can be used to calculate a predicted probability 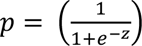, for being attributed to the severe PCC cluster as, where *z* = *predictor*_1_ × *coefficient*_1_ + ... + *predictor_q_* × *coefficient_q_* and *q* the number of selected predictors by the LASSO operator. Model performance was evaluated using the area under the receiver operating characteristic curve (ROC; AUC), and the optimal classification threshold was derived using the Youden index.

Statistical analysis and data curation were performed using R, Version 4.2.1 “Funny Looking Kid” (The R Foundation for Statistical Computing, Vienna, Austria) and Python 3, Version 3.12.5 (Python Software Foundation, Delaware, USA). The Hartigan-Wong algorithm was used for the k-means clustering, implemented in the *clusters* R package. The *glmnet* and the *caret* packages were used for the LASSO-penalized logistic regression and cross-validation.

### Ethics

The study adhered to the Declaration of Helsinki and all applicable regulations. The study protocol was approved by the *Ethikkommission der Fakultät für Medizin der Technischen Universität München* (Ethics Committee of the TUM University Hospital; approval 511/21, 2025-465-S-SB). Written informed consent was obtained from all participants aged 18–25 years; for participants aged 7–17 years, written informed consent was obtained from a parent or legal guardian, and age-appropriate assent was obtained from the participant. This observational registry study was registered at ClinicalTrials.gov (NCT05638724). No identifiable patient images or personal details are included in this Article; therefore, no additional publication permissions were required.

## RESULTS

### Patient Characteristics

120 CYP with PCC included 16/120 (13%) children (7-11 years), 71/12 (59%) adolescents (12-17 years), and 33/12 (28%) young adults (18-25 years). The percentage of females significantly increased with age (13% children, 62% adolescents, and 76% adults) (**Table 1**).

**Table 1.** Patient’s Basic Characteristics, Vaccination Status, and Symptoms During the Acute Phase of PCC-related SARS-CoV-2 Infection.

|  | **Overall<sup>a</sup>**<br>**N = 120** | **Children<sup>a</sup>**<br>**Age 7 to 11**<br>**years**<br>**N = 16** | **Adolescents<sup>a</sup>**<br>**Age 12 to 17**<br>**years**<br>**N = 71** | **Young Adults<sup>a</sup>**<br>**Age 18 to 25**<br>**years**<br>**N = 33** | **P-Value<sup>b</sup>** |
| --- | --- | --- | --- | --- | --- |
| Age<sup>a</sup>, years | 15·0 (13·0 - 18·0) | 11·0 (9·8 - 11·0) | 15·0 (13·0 - 16·0) | 21·0 (19·0 - 23·0) | NA |
| Sex, female | 71 / 120 (59%) | 2 / 16 (13%) | 44 / 71 (62%) | 25 / 33 (76%) | <0·001 |
| Body height<sup>a</sup>, cm | 167·0 (160·8 - 173·8) | 146·0 (141·8 - 152·3) | 167·0 (161·8 - 173·3) | 170·1 (166·5 - 177·0) | NA |
| Body weight<sup>a</sup>, kg | 58·5 (48·4 - 68·6) | 35·0 (31·7 - 45·8) | 60·7 (50·9 - 68·6) | 62·5 (56·6 - 75·0) | NA |
| Body mass index<sup>a</sup>, kg/m<sup>2</sup> | 20·7 (18·4 - 22·9) | 17·1 (15·9 - 19·0) | 21·3 (18·8 - 22·9) | 21·1 (19·5 - 24·7) | <0·001 |
| Allergies | 51 / 116 (44%) | 6 / 15 (40%) | 29 / 69 (42%) | 16 / 32 (50%) | 0·714 |
| Allergies since PCC onset | 4 / 39 (10%) | 1 / 2 (50%) | 2 / 22 (9·1%) | 1 / 15 (6·7%) | 0·214 |
| Susceptibility to infection<sup>c</sup> | 26 / 110 (24%) | 2 / 14 (14%) | 15 / 65 (23%) | 9 / 31 (29%) | 0·614 |
| Standard vaccination according to STIKO | 93 / 95 (98%) | 12 / 13 (92%) | 59 / 60 (98%) | 22 / 22 (100%) | 0·308 |
| COVID-19 vaccinations before symptom onset | NA | NA | NA | NA | 0·007 |
| none | 82 / 116 (71%) | 14 / 16 (88%) | 40 / 69 (58%) | 28 / 31 (90%) | NA |
| 1 | 0 / 116 (0%) | 0 / 16 (0%) | 0 / 69 (0%) | 0 / 31 (0%) | NA |
| 2 | 26 / 116 (22%) | 2 / 16 (13%) | 21 / 69 (30%) | 3 / 31 (10%) | NA |
| 3 | 8 / 116 (7%) | 0 / 16 (0%) | 8 / 69 (12%) | 0 / 31 (0%) | NA |
| PCC-triggering SARS-CoV-2 infection | NA | NA | NA | NA | 0·430 |
| 1st | 112 / 120 (93%) | 14 / 16 (88%) | 66 / 71 (93%) | 32 / 33 (97%) | NA |
| 2nd | 8 / 120 (7%) | 2 / 16 (13%) | 5 / 71 (7%) | 1 / 33 (3%) | NA |
| Documentation of SARS-CoV-2 infection | NA | NA | NA | NA | 0·166 |
| PCR | 106 / 120 (88%) | 14 / 16 (88%) | 61 / 71 (86%) | 31 / 33 (94%) | NA |
| Rapid antigen test | 8 / 120 (7%) | 2 / 16 (13%) | 6 / 71 (9%) | 0 / 33 (0%) | NA |
| Serology | 6 / 120 (5%) | 0 / 16 (0%) | 4 / 71 (6%) | 2 / 33 (6%) | NA |
| Time from infection to PCC symptom onset, days | 0.5 (-0.3 - 30.8) | 0.0 (-0.3 - 29.5) | 12.0 (.0 - 38.5) | 0.0 (-3.0 - 8.0) | 0.005 |
| Time from infection to diagnosis, months | 8.2 (5.4 - 12.4) | 6.5 (5.6 - 8.8) | 7.5 (5.0 - 12.1) | 10.8 (7.9 - 14.5) | 0.007 |
| Acute symptom treatment | NA | NA | NA | NA | 0.806 |
| No medical visit | 85 / 117 (73%) | 12 / 16 (75%) | 48 / 68 (71%) | 25 / 33 (76%) | NA |
| Outpatient | 23 / 117 (20%) | 4 / 16 (25%) | 13 / 68 (19%) | 6 / 33 (18%) | NA |
| Hospitalized | 9 / 117 (8%) | 0 / 16 (0%) | 7 / 68 (10%) | 2 / 33 (6%) | NA |
| Number of acute symptoms reported | 16.0 (11.0 - 21.0) | 17.0 (13.0 - 18.5) | 14.5 (11.0 - 20.0) | 16.0 (12.0 - 23.0) | 0.580 |
| **Acute Symptoms** | NA | NA | NA | NA | NA |
| Fatigue | 104 / 116 (90%) | 15 / 15 (100%) | 60 / 68 (88%) | 29 / 33 (88%) | 0.503 |
| Headaches | 103 / 116 (89%) | 13 / 15 (87%) | 60 / 68 (88%) | 30 / 33 (91%) | 0.918 |
| Sore throat | 91 / 116 (78%) | 14 / 15 (93%) | 55 / 68 (81%) | 22 / 33 (67%) | 0.107 |
| Persistent dry cough | 87 / 116 (75%) | 11 / 15 (73%) | 54 / 68 (79%) | 22 / 33 (67%) | 0.375 |
| Trouble in concentrating | 85 / 116 (73%) | 13 / 15 (87%) | 47 / 68 (69%) | 25 / 33 (76%) | 0.415 |
| Shortness of breath | 84 / 116 (72%) | 11 / 15 (73%) | 50 / 68 (74%) | 23 / 33 (70%) | 0.955 |
| Cold | 83 / 116 (72%) | 9 / 15 (60%) | 53 / 68 (78%) | 21 / 33 (64%) | 0.184 |
| Dizziness | 75 / 116 (65%) | 9 / 15 (60%) | 43 / 68 (63%) | 23 / 33 (70%) | 0.752 |
| Muscle pain | 70 / 116 (60%) | 8 / 15 (53%) | 43 / 68 (63%) | 19 / 33 (58%) | 0.722 |
| Fever | 65 / 116 (56%) | 11 / 15 (73%) | 38 / 68 (56%) | 16 / 33 (48%) | 0.274 |
| Loss of appetite | 61 / 116 (53%) | 13 / 15 (87%) | 33 / 68 (49%) | 15 / 33 (45%) | 0.017 |
| Chest tightness | 59 / 116 (51%) | 9 / 15 (60%) | 30 / 68 (44%) | 20 / 33 (61%) | 0.224 |
| Joint pain | 59 / 116 (51%) | 7 / 15 (47%) | 34 / 68 (50%) | 18 / 33 (55%) | 0.859 |
| Nausea | 53 / 116 (46%) | 8 / 15 (53%) | 34 / 68 (50%) | 11 / 33 (33%) | 0.235 |
| Stomach pain | 51 / 116 (44%) | 10 / 15 (67%) | 28 / 68 (41%) | 13 / 33 (39%) | 0.163 |
| Pain on breathing | 47 / 116 (41%) | 6 / 15 (40%) | 24 / 68 (35%) | 17 / 33 (52%) | 0.297 |
| Palpitations | 45 / 116 (39%) | 6 / 15 (40%) | 23 / 68 (34%) | 16 / 33 (48%) | 0.364 |
| Tachykardia | 45 / 116 (39%) | 4 / 15 (27%) | 22 / 68 (32%) | 19 / 33 (58%) | 0.030 |
| Swollen lymph nodes | 44 / 116 (38%) | 8 / 15 (53%) | 27 / 68 (40%) | 9 / 33 (27%) | 0.202 |
| Loss of smell | 44 / 116 (38%) | 0 / 15 (0%) | 25 / 68 (37%) | 19 / 33 (58%) | <0.001 |
| Apathy | 43 / 116 (37%) | 5 / 15 (33%) | 22 / 68 (32%) | 16 / 33 (48%) | 0.275 |
| Light-headedness | 42 / 116 (36%) | 4 / 15 (27%) | 24 / 68 (35%) | 14 / 33 (42%) | 0.558 |
| Weight loss | 38 / 116 (33%) | 5 / 15 (33%) | 21 / 68 (31%) | 12 / 33 (36%) | 0.883 |
| Loss of taste | 38 / 116 (33%) | 0 / 15 (0%) | 23 / 68 (34%) | 15 / 33 (45%) | 0.003 |
| Altered taste | 32 / 116 (28%) | 4 / 15 (27%) | 19 / 68 (28%) | 9 / 33 (27%) | >0.999 |
| Altered smell | 29 / 116 (25%) | 3 / 15 (20%) | 17 / 68 (25%) | 9 / 33 (27%) | 0.909 |
| Breath-independent chest pain | 28 / 116 (24%) | 6 / 15 (40%) | 12 / 68 (18%) | 10 / 33 (30%) | 0.100 |
| Numbness or tingling | 26 / 116 (22%) | 5 / 15 (33%) | 14 / 68 (21%) | 7 / 33 (21%) | 0.561 |
| Diarrhoea | 25 / 116 (22%) | 9 / 15 (60%) | 9 / 68 (13%) | 7 / 33 (21%) | <0.001 |
| Fainting/blackouts | 14 / 116 (12%) | 1 / 15 (7%) | 11 / 68 (16%) | 2 / 33 (6%) | 0.393 |
| Vomiting | 13 / 116 (11%) | 2 / 15 (13%) | 7 / 68 (10%) | 4 / 33 (12%) | 0.841 |
| Conjunctivitis | 11 / 116 (10%) | 0 / 15 (0%) | 7 / 68 (10%) | 4 / 33 (12%) | 0.483 |
| Skin rash | 9 / 116 (8%) | 2 / 15 (13%) | 3 / 68 (4%) | 4 / 33 (12%) | 0.226 |
| Pain in the limbs | 7 / 116 (6%) | 0 / 15 (0%) | 3 / 68 (4%) | 4 / 33 (12%) | 0.252 |
| COVID-19 toes (purple, pink, bluish) | 6 / 116 (5%) | 0 / 15 (0%) | 5 / 68 (7%) | 1 / 33 (3%) | 0.585 |

**a** Data are expressed as n/N (%), median (IQR), or mean (SD), as appropriate. Percentages may not total 100% because of rounding.
**b** P-values were calculated using the $\chi^2$ test, Fisher exact test, or Kruskal-Wallis test, as appropriate.
**c** Susceptibility to infection was defined as $\geq 1$ of the following: recurrent infections at the same site, >2 antibiotic courses/year, hospitalization due to infection, $\geq 4$ episodes of herpes labialis/year, at least one episode of genital herpes or herpes zoster.
**d** Abbreviations: PCR, polymerase chain reaction; SARS-CoV-2, severe acute respiratory syndrome coronavirus 2; STIKO, Standing Committee on Vaccination at the Robert Koch Institute (Germany).
NA = not available.

### Characteristics of the Acute Phase of the SARS-CoV-2 Infection

Documented or probable SARS-CoV-2 infections occurred between February 2020 and January 2023, spanning pandemic waves in Germany with dominant Alpha (43/114 (38%)), Delta (25/114 (22%)), and Omicron variants (46/114 (40%)).

112/120 (93%) and 8/120 (7%) patients reported their first or second SARS-CoV-2 infection as PCC trigger, respectively. The triggering SARS-CoV-2 infection was indicated by PCR in 106/120 (88%), rapid antigen testing in 8/120 (7%), or serology together with contact in 6/120 (5%) CYP. 82/116 (71%) were not COVID-19 vaccinated prior to the index infection. A triggering breakthrough infection was reported in 34/116 (29%) patients (**Table 1**).

All patients remembered a median number of 16 (IQR: 11-21) acute COVID-19 symptoms during the index infection, with no significant difference between the three age groups (P = 0·579). The five most frequently reported COVID-19 symptoms were fatigue, headaches, sore throat, persistent dry cough, and trouble concentrating (**Table 1**). Loss of appetite (P = 0·017) and diarrhea (P < 0·001) were significantly more frequent in children. Heart racing (P = 0·030), loss of smell (P < 0·001) of taste (P = 0·003) were significantly more frequent in adolescents and young adults. Only 9/117 (8%) patients were hospitalized for their index infection (**Table 1**).

### Characteristics of Post-COVID-Condition

Time from acute symptom onset to diagnosis was 8·2 month in median (IQR: 5·4 - 12·4) and significantly greater for young adults compare to children and adolescents (P = 0·007). Patients presented with various PCC-associated medical diagnoses (**Suppl. Table S1, Appendix p.2**). The three most frequent were fatigue (ICD-10-GM R53) (81/120, 68%), brain fog (ICD-10-GM F06.7) (64/120, 53%), and headaches (ICD-10-GM R51) (49/120, 41%). Brain fog (ICD-10-GM F06.7) was significantly more frequent in young adults compared to children and adolescents (P = 0·034).

The median number of PCC symptoms at the time of diagnosis was 29 (IQR: 21 – 37) and the most frequent symptoms were fatigue, exercise intolerance, reduced endurance, troubles in concentrating, and headaches, as indicated by the MLCSQ (**Table 2**). The median FSS score was 6·4 (IQR 5·8 to 6·6), reflecting pronounced fatigue (**Table 3**). The DSQ-PEM indicated PEM in 112/119 (94%) patients, with a duration of 14-23 or ≥24 hours reported by 19/114 (17%) and 23/114 (20%) patients, respectively (**Table 3**). The COMPASS 31 revealed a median score for OI of 20·0 (12·0 - 28·0) and a median total score of 27·8 (17·8 - 38·8), reflecting severe autonomic distress (**Table 3**).

**Table 2.** Number of Chronic Postviral Symptoms Assessed by the Munich Long COVID Symptom Questionnaire (MLCSQ).

|  | **Overall<sup>a</sup>** | **Children<sup>a</sup>** | **Adolescents<sup>a</sup>** | **Young Adults<sup>a</sup>** | **P-Value<sup>b</sup>** |
| --- | --- | --- | --- | --- | --- |
|  | **N = 120** | **Age 7 to 11**<br>**years**<br>**N = 16** | **Age 12 to 17**<br>**years**<br>**N = 71** | **Age 18 to 25**<br>**years**<br>**N = 33** |  |
| Number of 83 possible PCC symptoms reported | 29.0 (21.0 - 37.0) | 29.5 (18.0 - 35.25) | 29.0 (20.0 - 36.0) | 31.0 (24.5 - 37.75) | 0.448 |
| **Fatigue / Exercise intolerance** | NA | NA | NA | NA | NA |
| Fatigue | 105 / 109 (96%) | 14 / 16 (88%) | 63 / 65 (97%) | 28 / 28 (100%) | 0.097 |
| Worsening of symptoms following mild mental and/or physical activity (exercise intolerance) | 104 / 109 (95%) | 14 / 16 (88%) | 62 / 65 (95%) | 28 / 28 (100%) | 0.162 |
| Reduced endurance | 103 / 109 (94%) | 14 / 16 (88%) | 61 / 65 (94%) | 28 / 28 (100%) | 0.203 |
| **Neurological** | NA | NA | NA | NA | NA |
| Trouble in concentrating | 97 / 109 (89%) | 15 / 16 (94%) | 55 / 65 (85%) | 27 / 28 (96%) | 0.200 |
| Headaches | 96 / 109 (88%) | 14 / 16 (88%) | 56 / 65 (86%) | 26 / 28 (93%) | 0.656 |
| Dizziness | 84 / 109 (77%) | 10 / 16 (62%) | 54 / 65 (83%) | 20 / 28 (71%) | 0.153 |
| Forgetfulness | 81 / 109 (74%) | 11 / 16 (69%) | 47 / 65 (72%) | 23 / 28 (82%) | 0.523 |
| Light-headedness | 54 / 109 (50%) | 3 / 16 (19%) | 33 / 65 (51%) | 18 / 28 (64%) | 0.014 |
| Problems with balance | 50 / 109 (46%) | 6 / 16 (38%) | 33 / 65 (51%) | 11 / 28 (39%) | 0.456 |
| Numbness or tingling | 34 / 109 (31%) | 7 / 16 (44%) | 17 / 65 (26%) | 10 / 28 (36%) | 0.331 |
| Slowness of movement | 28 / 109 (26%) | 5 / 16 (31%) | 16 / 65 (25%) | 7 / 28 (25%) | 0.858 |
| Problems with gait/falls | 21 / 109 (19%) | 4 / 16 (25%) | 16 / 65 (25%) | 1 / 28 (4%) | 0.051 |
| Seizures | 8 / 109 (7%) | 1 / 16 (6%) | 6 / 65 (9%) | 1 / 28 (4%) | 0.621 |
| Can't feel one side of body or face | 7 / 109 (6%) | 0 / 16 (0%) | 6 / 65 (9%) | 1 / 28 (4%) | 0.312 |
| Can't move one side of body or face | 1 / 109 (1%) | 0 / 16 (0%) | 0 / 65 (0%) | 1 / 28 (4%) | 0.232 |
| **Pulmonary** | NA | NA | NA | NA | NA |
| Shortness of breath with activity | 88 / 109 (81%) | 12 / 16 (75%) | 54 / 65 (83%) | 22 / 28 (79%) | 0.722 |
| Persistent dry cough | 48 / 109 (44%) | 7 / 16 (44%) | 30 / 65 (46%) | 11 / 28 (39%) | 0.829 |
| Shortness of breath at rest | 44 / 109 (40%) | 4 / 16 (25%) | 30 / 65 (46%) | 10 / 28 (36%) | 0.256 |
| Pain on breathing | 43 / 109 (39%) | 3 / 16 (19%) | 24 / 65 (37%) | 16 / 28 (57%) | 0.035 |
| Congestion | 36 / 109 (33%) | 2 / 16 (12%) | 24 / 65 (37%) | 10 / 28 (36%) | 0.167 |
| **Cardiovascular** | NA | NA | NA | NA | NA |
| Orthostatic intolerance | 83 / 109 (76%) | 6 / 16 (38%) | 53 / 65 (82%) | 24 / 28 (86%) | <0.001 |
| Heart pounding | 55 / 109 (50%) | 6 / 16 (38%) | 31 / 65 (48%) | 18 / 28 (64%) | 0.181 |
| Chest tightness | 54 / 109 (50%) | 4 / 16 (25%) | 30 / 65 (46%) | 20 / 28 (71%) | 0.009 |
| Heart racing | 51 / 109 (47%) | 5 / 16 (31%) | 30 / 65 (46%) | 16 / 28 (57%) | 0.251 |
| Breath-independent chest pain | 25 / 109 (23%) | 3 / 16 (19%) | 13 / 65 (20%) | 9 / 28 (32%) | 0.403 |
| Fainting and/or Blackouts | 16 / 109 (15%) | 1 / 16 (6%) | 15 / 65 (23%) | 0 / 28 (0%) | 0.009 |
| **Sleep** | NA | NA | NA | NA | NA |
| Sleeping more | 81 / 109 (74%) | 10 / 16 (62%) | 47 / 65 (72%) | 24 / 28 (86%) | 0.201 |
| Other sleep disturbances | 50 / 109 (46%) | 7 / 16 (44%) | 30 / 65 (46%) | 13 / 28 (46%) | 0.983 |
| Sleeping less | 37 / 109 (34%) | 3 / 16 (19%) | 22 / 65 (34%) | 12 / 28 (43%) | 0.267 |
| **Mental health** | NA | NA | NA | NA | NA |
| Listlessness / Lack of motivation | 80 / 109 (73%) | 11 / 16 (69%) | 50 / 65 (77%) | 19 / 28 (68%) | 0.597 |
| Loss of interest/pleasure | 61 / 109 (56%) | 7 / 16 (44%) | 37 / 65 (57%) | 17 / 28 (61%) | 0.535 |
| Depressed mood | 59 / 109 (54%) | 5 / 16 (31%) | 37 / 65 (57%) | 17 / 28 (61%) | 0.131 |
| Anxiety | 44 / 109 (40%) | 7 / 16 (44%) | 25 / 65 (38%) | 12 / 28 (43%) | 0.884 |
| Behavioural change | 35 / 109 (32%) | 5 / 16 (31%) | 20 / 65 (31%) | 10 / 28 (36%) | 0.893 |
| Tic | 11 / 109 (10%) | 2 / 16 (12%) | 8 / 65 (12%) | 1 / 28 (4%) | 0.414 |
| Hallucinations | 6 / 109 (6%) | 1 / 16 (6%) | 5 / 65 (8%) | 0 / 28 (0%) | 0.325 |
| **Musculoskeletal** | NA | NA | NA | NA | NA |
| Muscle pain | 69 / 109 (63%) | 12 / 16 (75%) | 38 / 65 (58%) | 19 / 28 (68%) | 0.397 |
| Muscle weakness | 63 / 109 (58%) | 9 / 16 (56%) | 35 / 65 (54%) | 19 / 28 (68%) | 0.451 |
| Back pain | 59 / 109 (54%) | 9 / 16 (56%) | 33 / 65 (51%) | 17 / 28 (61%) | 0.666 |
| Joint pain | 52 / 109 (48%) | 9 / 16 (56%) | 29 / 65 (45%) | 14 / 28 (50%) | 0.678 |
| Muscle twitching | 36 / 109 (33%) | 4 / 16 (25%) | 21 / 65 (32%) | 11 / 28 (39%) | 0.614 |
| Muscle tremor | 34 / 109 (31%) | 3 / 16 (19%) | 23 / 65 (35%) | 8 / 28 (29%) | 0.411 |
| Muscle stiffness | 17 / 109 (16%) | 3 / 16 (19%) | 7 / 65 (11%) | 7 / 28 (25%) | 0.207 |
| Joint swelling | 5 / 109 (5%) | 2 / 16 (12%) | 2 / 65 (3%) | 1 / 28 (4%) | 0.260 |
| Redness and/or overheating of joints | 4 / 109 (4%) | 0 / 16 (0%) | 3 / 65 (5%) | 1 / 28 (4%) | 0.679 |
| **Gastro-intestinal** | NA | NA | NA | NA | NA |
| Stomach pain | 65 / 109 (60%) | 10 / 16 (62%) | 37 / 65 (57%) | 18 / 28 (64%) | 0.777 |
| Nausea | 55 / 109 (50%) | 10 / 16 (62%) | 30 / 65 (46%) | 15 / 28 (54%) | 0.468 |
| Loss of appetite | 48 / 109 (44%) | 11 / 16 (69%) | 25 / 65 (38%) | 12 / 28 (43%) | 0.091 |
| Diarrhoea | 39 / 109 (36%) | 7 / 16 (44%) | 20 / 65 (31%) | 12 / 28 (43%) | 0.414 |
| Flatulence | 38 / 109 (35%) | 8 / 16 (50%) | 19 / 65 (29%) | 11 / 28 (39%) | 0.251 |
| Bloatedness | 29 / 109 (27%) | 4 / 16 (25%) | 13 / 65 (20%) | 12 / 28 (43%) | 0.072 |
| Constipation | 25 / 109 (23%) | 5 / 16 (31%) | 10 / 65 (15%) | 10 / 28 (36%) | 0.070 |
| Weight loss | 16 / 109 (15%) | 2 / 16 (12%) | 9 / 65 (14%) | 5 / 28 (18%) | 0.851 |
| Vomiting | 6 / 109 (6%) | 1 / 16 (6%) | 4 / 65 (6%) | 1 / 28 (4%) | 0.873 |
| **Body Temperature** | NA | NA | NA | NA | NA |
| Increased sweating | 63 / 109 (58%) | 11 / 16 (69%) | 39 / 65 (60%) | 13 / 28 (46%) | 0.301 |
| Chills | 34 / 109 (31%) | 6 / 16 (38%) | 18 / 65 (28%) | 10 / 28 (36%) | 0.627 |
| Fever > 38.0 °C | 18 / 109 (16%) | 5 / 16 (31%) | 8 / 65 (12%) | 5 / 28 (18%) | 0.184 |
| **Ears-Nose-Throat (ENT)** | NA | NA | NA | NA | NA |
| Cold | 46 / 109 (42%) | 6 / 16 (38%) | 29 / 65 (45%) | 11 / 28 (39%) | 0.820 |
| Sore throat | 40 / 109 (37%) | 7 / 16 (44%) | 24 / 65 (37%) | 9 / 28 (32%) | 0.743 |
| Swollen lymph nodes | 27 / 109 (25%) | 6 / 16 (38%) | 14 / 65 (22%) | 7 / 28 (25%) | 0.416 |
| Dryness of the mouth | 27 / 109 (25%) | 2 / 16 (12%) | 17 / 65 (26%) | 8 / 28 (29%) | 0.455 |
| Altered taste | 24 / 109 (22%) | 5 / 16 (31%) | 10 / 65 (15%) | 9 / 28 (32%) | 0.127 |
| Problem swallowing | 24 / 109 (22%) | 5 / 16 (31%) | 14 / 65 (22%) | 5 / 28 (18%) | 0.581 |
| Ringing in ears | 24 / 109 (22%) | 4 / 16 (25%) | 15 / 65 (23%) | 5 / 28 (18%) | 0.816 |
| Altered smell | 22 / 109 (20%) | 3 / 16 (19%) | 10 / 65 (15%) | 9 / 28 (32%) | 0.179 |
| Ear pain | 22 / 109 (20%) | 3 / 16 (19%) | 15 / 65 (23%) | 4 / 28 (14%) | 0.618 |
| Sinus disorder | 20 / 109 (18%) | 1 / 16 (6%) | 13 / 65 (20%) | 6 / 28 (21%) | 0.395 |
| Voice disorder | 18 / 109 (16%) | 2 / 16 (12%) | 12 / 65 (18%) | 4 / 28 (14%) | 0.792 |
| Loss of smell | 14 / 109 (13%) | 0 / 16 (0%) | 8 / 65 (12%) | 6 / 28 (21%) | 0.121 |
| Hearing impairment | 12 / 109 (11%) | 3 / 16 (19%) | 7 / 65 (11%) | 2 / 28 (7%) | 0.494 |
| Loss of taste | 9 / 109 (8%) | 1 / 16 (6%) | 4 / 65 (6%) | 4 / 28 (14%) | 0.405 |
| **Ophthalmic** | NA | NA | NA | NA | NA |
| Light sensitivity | 44 / 109 (40%) | 7 / 16 (44%) | 28 / 65 (43%) | 9 / 28 (32%) | 0.588 |
| Dry eyes | 29 / 109 (27%) | 2 / 16 (12%) | 17 / 65 (26%) | 10 / 28 (36%) | 0.243 |
| Vision impairment | 24 / 109 (22%) | 5 / 16 (31%) | 14 / 65 (22%) | 5 / 28 (18%) | 0.581 |
| Conjunctivitis | 24 / 109 (22%) | 4 / 16 (25%) | 13 / 65 (20%) | 7 / 28 (25%) | 0·826 |
| **Urogenital** | NA | NA | NA | NA | NA |
| Painful menstruation | 29 / 109 (27%) | 0 / 16 (0%) | 19 / 65 (29%) | 10 / 28 (36%) | 0·027 |
| Problems passing urine | 7 / 109 (6%) | 1 / 16 (6%) | 2 / 65 (3%) | 4 / 28 (14%) | 0·129 |
| Changes in sexual function | 1 / 109 (1%) | 0 / 16 (0%) | 0 / 65 (0%) | 1 / 28 (4%) | 0·232 |
| **Dermatological** | NA | NA | NA | NA | NA |
| Hair loss | 26 / 109 (24%) | 0 / 16 (0%) | 14 / 65 (22%) | 12 / 28 (43%) | 0·005 |
| Skin rash | 19 / 109 (17%) | 3 / 16 (19%) | 8 / 65 (12%) | 8 / 28 (29%) | 0·164 |
| COVID-19 toes (purple, pink, bluish) | 14 / 109 (13%) | 0 / 16 (0%) | 10 / 65 (15%) | 4 / 28 (14%) | 0·248 |
| Other changes in skin color on hands and/or feet (red, white) | 14 / 109 (13%) | 2 / 16 (12%) | 6 / 65 (9%) | 6 / 28 (21%) | 0·272 |
| Swollen ankles / oedema | 6 / 109 (6%) | 0 / 16 (0%) | 3 / 65 (5%) | 3 / 28 (11%) | 0·288 |

**a** Data are expressed as n/N (%) or median (IQR), as appropriate. Percentages may not total 100% because of rounding.
**b** P values were calculated using the $\chi^2$ test or Kruskal-Wallis test, as appropriate.
**c** Abbreviations: MLCSQ, Munich Long COVID Symptom Questionnaire, PCC, Post-COVID Condition
NA = not available.

**Table 3.**
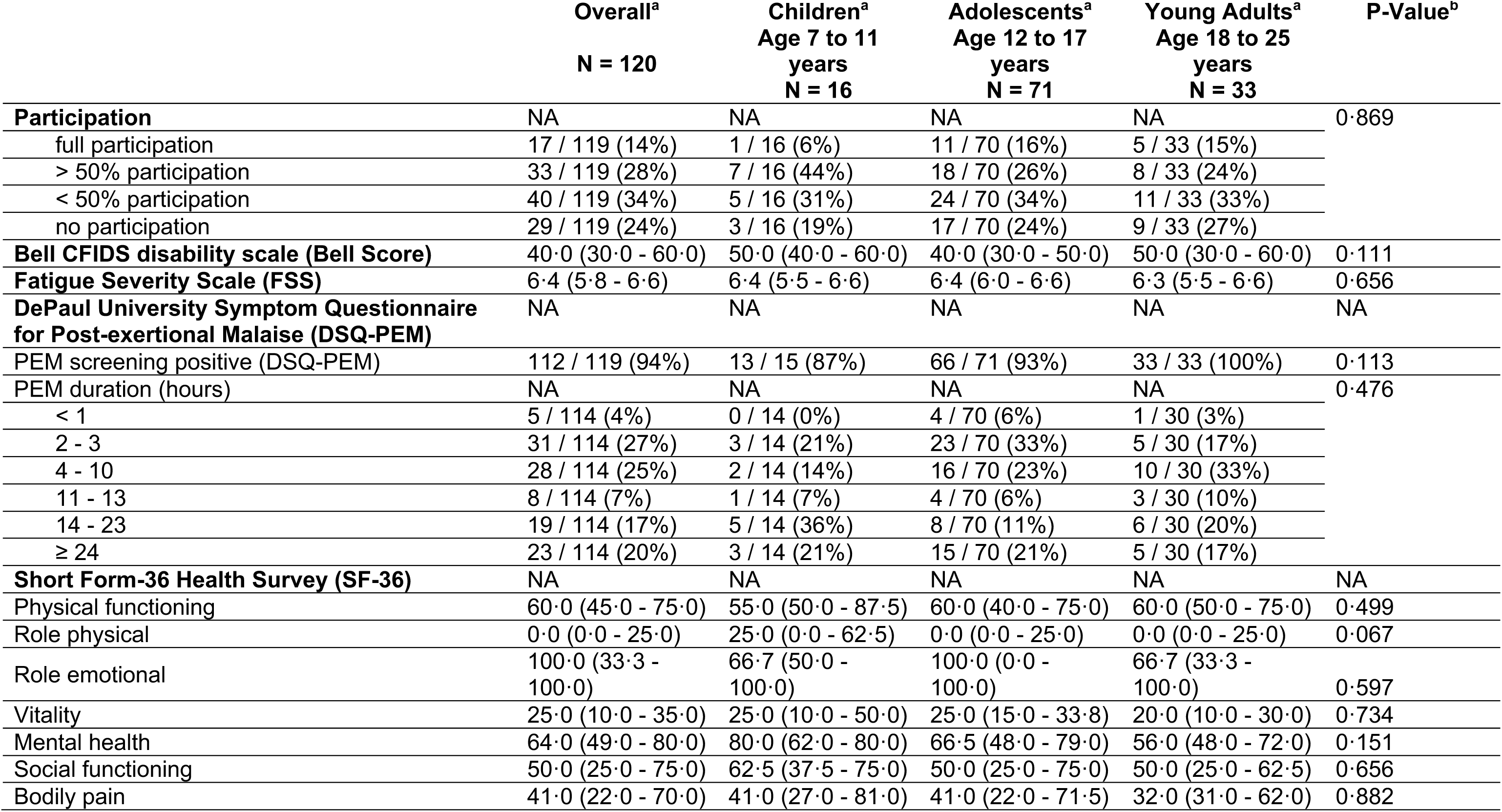
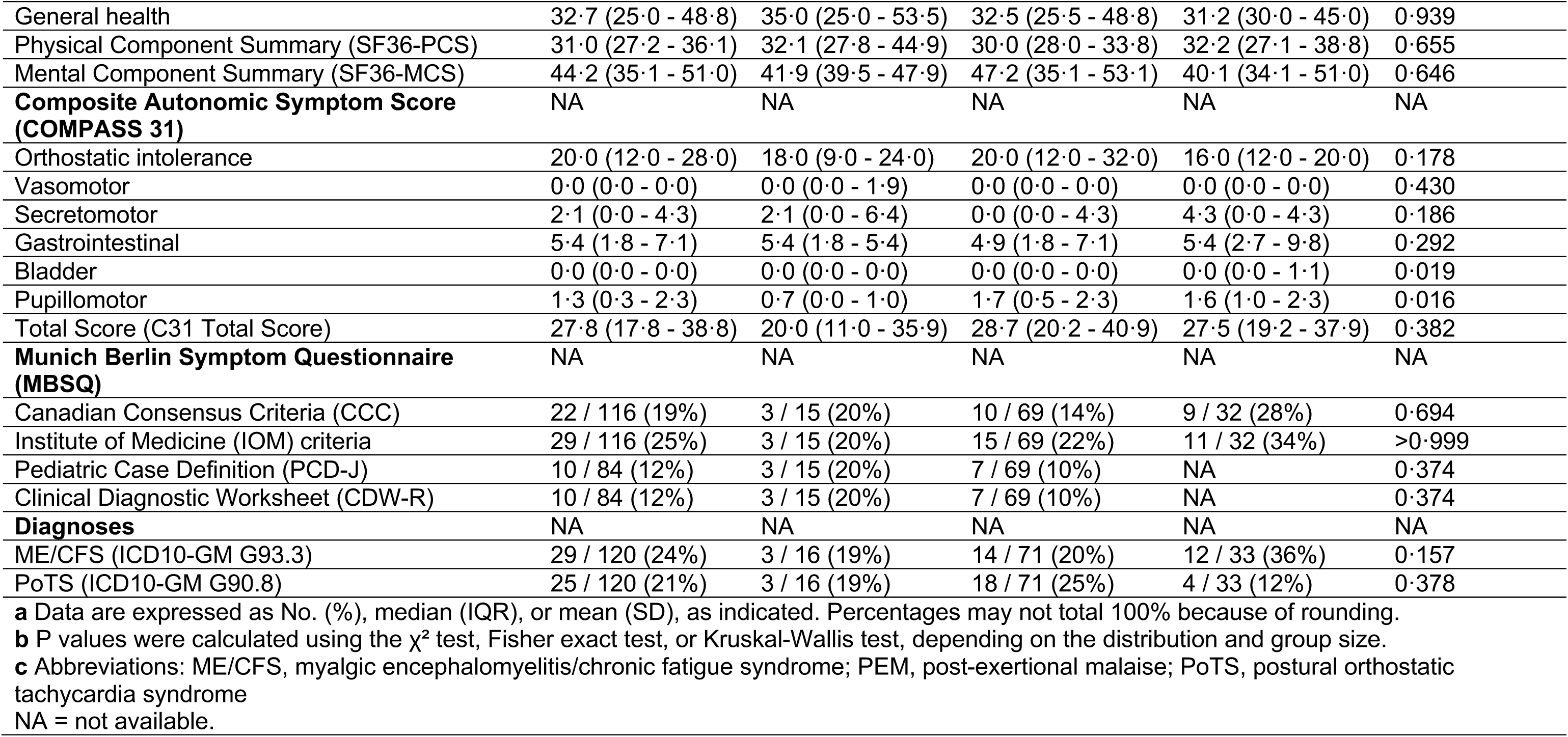
Characteristics of Post-COVID Condition Assessed by Questionnaires and Defined by Specific ICD-10 Diagnoses.

The median Bell Score was 40% (IQR 30 to 60), indicating significant impairment of daily life (**Table 3**). 40/119 (34%) and 29/119 (24%) CYP stated to have less than 50% or no participation in school or work, respectively (**Table 3**). SF-36 median scores were markedly reduced in all eight dimensions compared to a German normative sample (aged 14–20 years, N = 228), with a median PCS of 31·0 (27·2 - 36·1) versus 57·1 (54·6 – 58·8) (P < 0·025) and a median MCS of 44·2 (35·1 - 51·0) versus 53·5 (50·4 – 55·9) (P < 0·114) (**Table 3**). PoTS (ICD-10-GM G90·80) was diagnosed in 25/120 (21%) patients (**Table 3**).

### Sub-Group Comparison of ME/CFS vs. non-ME/CFS Patients

The diagnosis of ME/CFS (ICD-10-GM G93.3) was confirmed or probable after medical assessment in 29/120 (24·2%) patients (**Table 3**) and was more frequent with older age (children 19%, adolescents 20%, adults 36%).

Confirmed or probable ME/CFS compared to non-MECFS patients had a greater number of acute COVID-19 symptoms (median 17·5 (14·2 – 20·0) versus 14·0 (10·0 – 18·0); P = 0·021), a greater number of PCC symptoms (median 18·0 (16·0 – 22·0) versus 16·0 (11·5 – 20·0); P = 0·032), higher FSS scores (median 6·6 (6·3 – 6·8) versus 6·2 (5·7 – 6·6); P = 0·012), higher proportion of PEM duration ≥ 24h: 37·9% versus 14·0%, higher COMPASS 31 total scores (median 34·9 (25·6 - 43·5) versus 25·6 (16·8 - 36·5); P = 0·032), lower Bell Scores (median 30·0 (30·0 - 50·0) versus 50·0 (40·0 - 60·0); P < 0·001), and lower scores of SF36-PCS (median 27·1 (23·6 - 32·0) versus 31·5 (28·3 - 38·6); P = 0·025) and SF36-MCS (median 42·4 (29·5 - 47·9) versus 45·6 (36·0 - 52·7), P = 0·114). (**Suppl. Table S1, Appendix p.2**)

### Risk Factors Analysis Using Distinct Clinical Outcomes

Possible associations of individual candidate predictor variables during the acute phase of the SARS-CoV-2 infection and distinct outcomes at the time of PCC diagnoses were investigated: *i) Acute OI* was significantly associated with a higher FSS score (adj. coefficient: 0·377, P = 0·042), a higher COMPASS 31 total score (adj. coefficient: 12·570, P < 0·001), and a lower SF36-MCS (adj. coefficient: -6·185, P = 0·008); *ii)* the *presence of OI or GI symptoms* was significantly associated with a higher number of PCC symptoms (OI: adj. coefficient: 9·276, P < 0·001; GI: adj. coefficient: 9·668, P < 0·001); *iii) hospitalization due to COVID-19* was associated with a lower Bell score (adj. coefficient: -8·991, P = 0·176) and a lower SF36-MCS (adj. coefficient: -7·685, P = 0·048); *iv) a higher number of COVID-19 symptoms* was significantly associated with higher FSS scores (adj. coefficient: 0·045, P < 0·001), a higher COMPASS 31 total score (adj. coefficient: 0·633, P = 0·010), a lower SF36-PCS (adj. coefficient: -0·522, P = 0·001), a lower SF36-MCS (adj. coefficient: -0·335, P = 0·060), and a higher number of chronic symptoms (adj. coefficient: 1·142, P < 0·001); *v) acute trouble concentrating* were significantly associated with a higher FSS score (adj. coefficient: 0·502, P = 0·013), a higher SF36-PCS (adj. coefficient: -6·232, P = 0·007), a higher number of chronic symptoms (adj. coefficient: 10·9, P < 0·001) and a higher likelihood for a positive DSQ-PEM screening (adj. OR: 2·360, P = 0·019); *vi) older age* was significantly associated with a higher likelihood for positive PEM as indicated by the DSQ-PEM (adj. OR: 1·509, P = 0·009), independent of sex, and *vii) being female* was associated with a significantly higher likelihood of confirmed or suspected medical ME/CFS diagnosis (G93.3) and a significantly higher COMPASS 31 total score (adj. coefficient: 9·657, P = 0·005). None of the acute features were significantly associated with brain fog (F06.7). Results are summarized in **Table 4**.

**Table 4.**
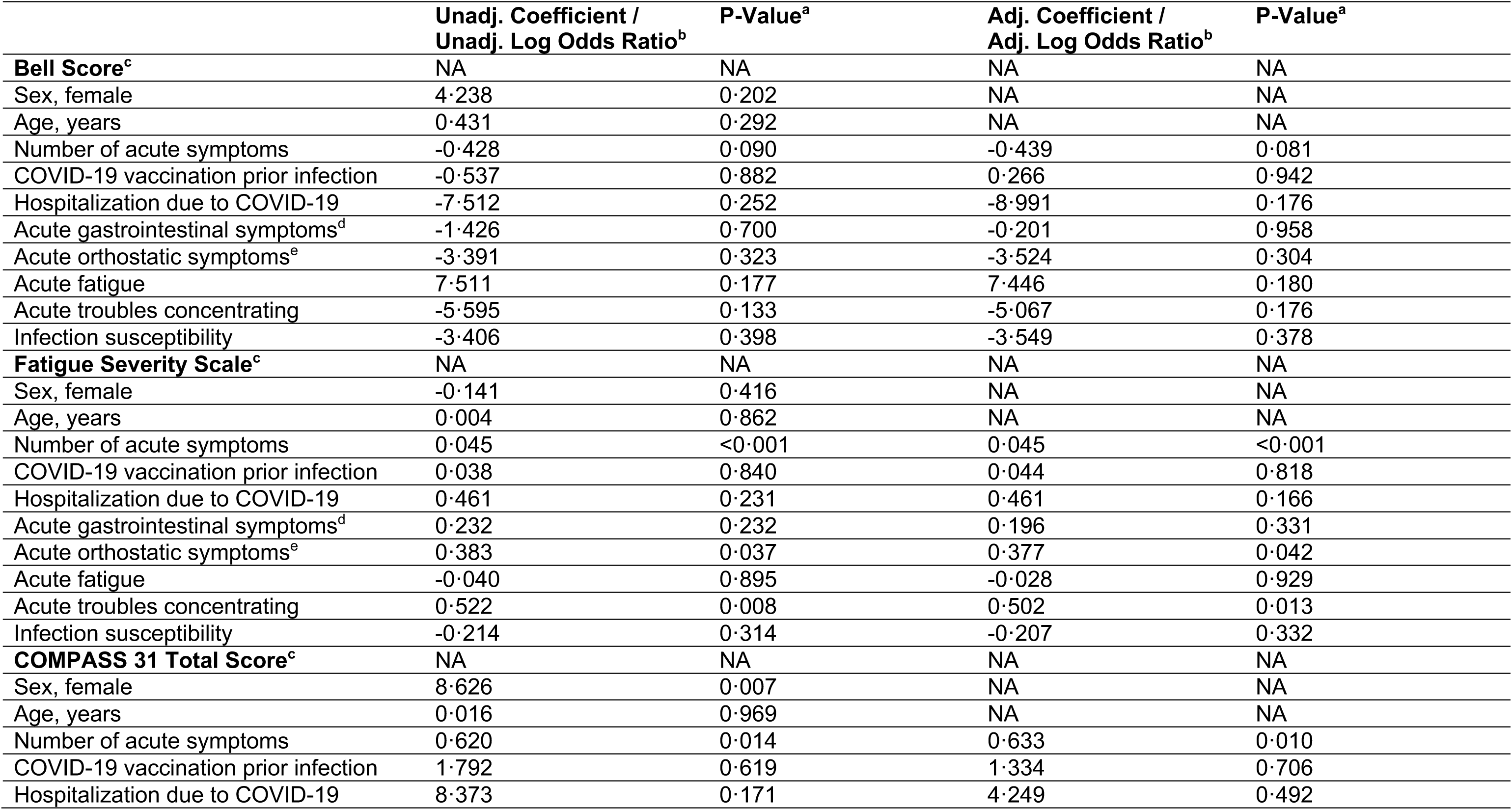
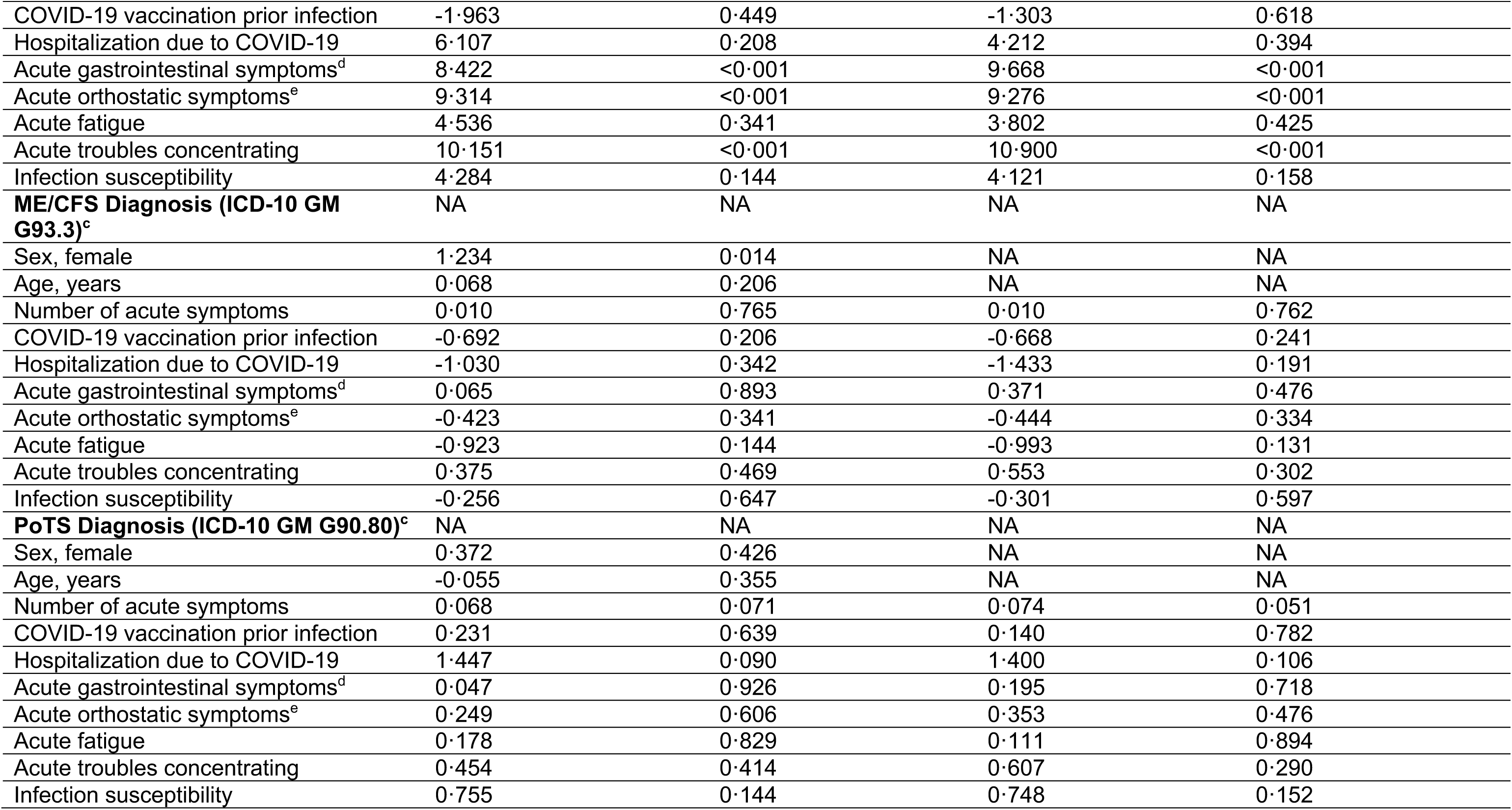
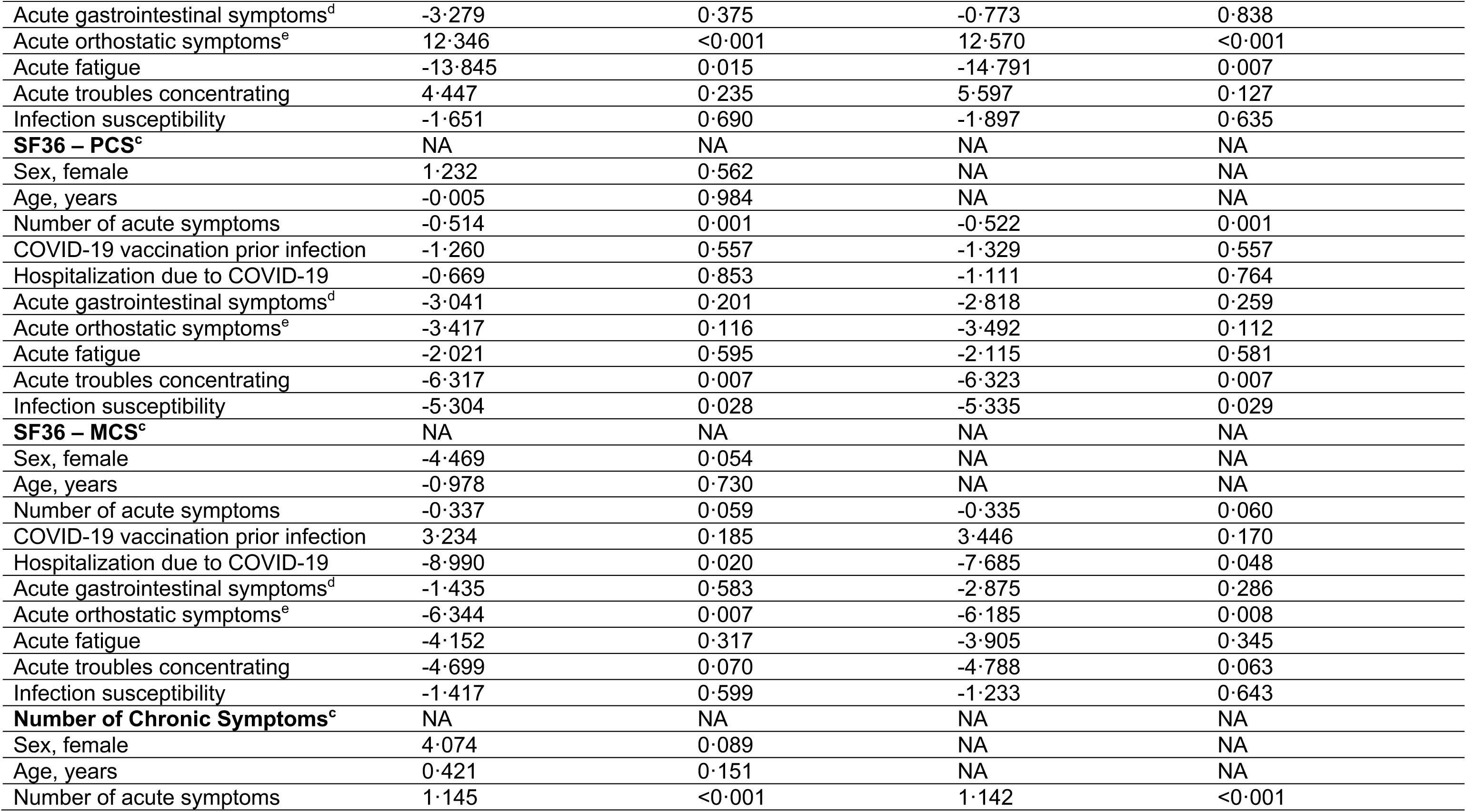
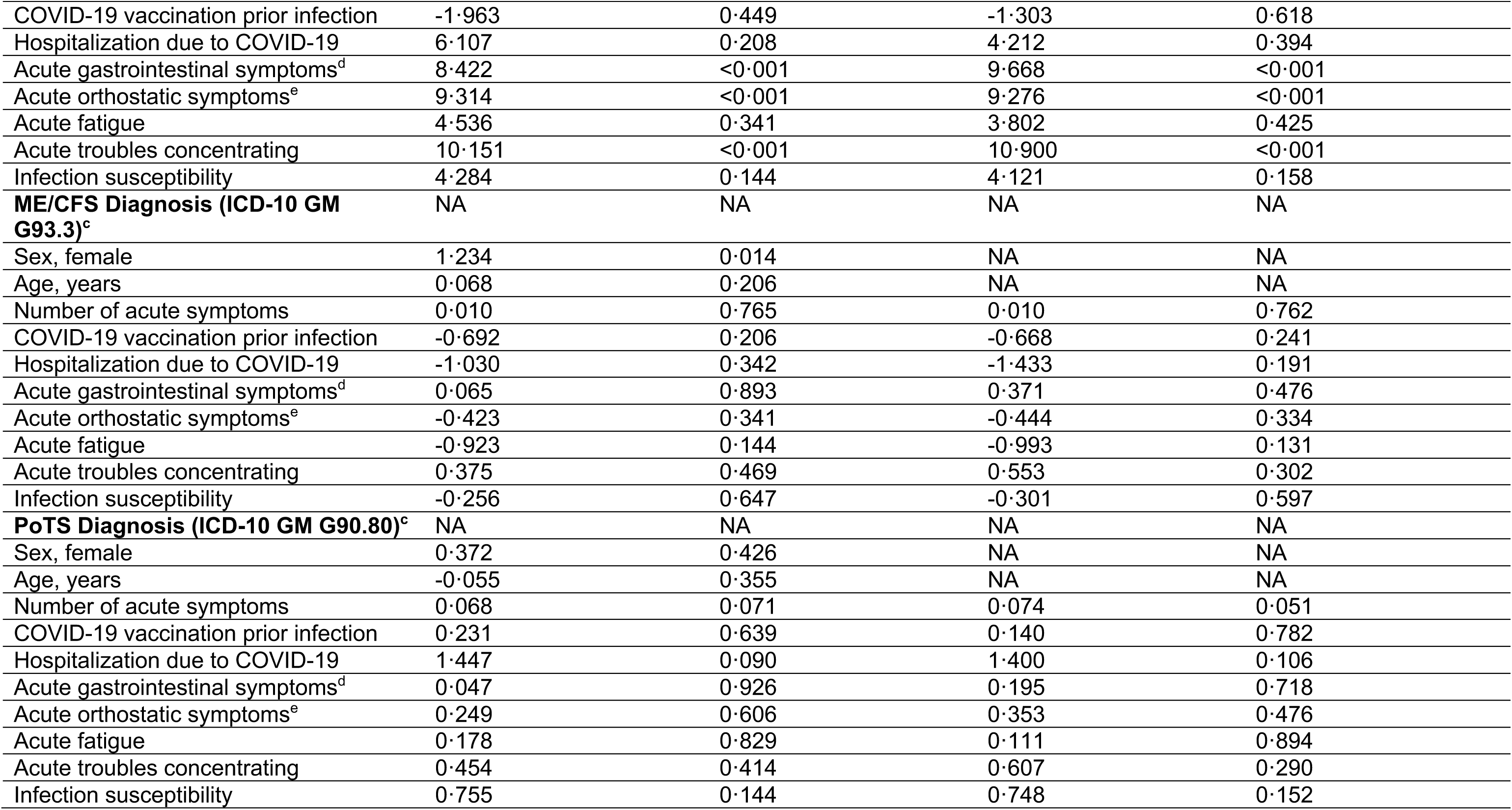
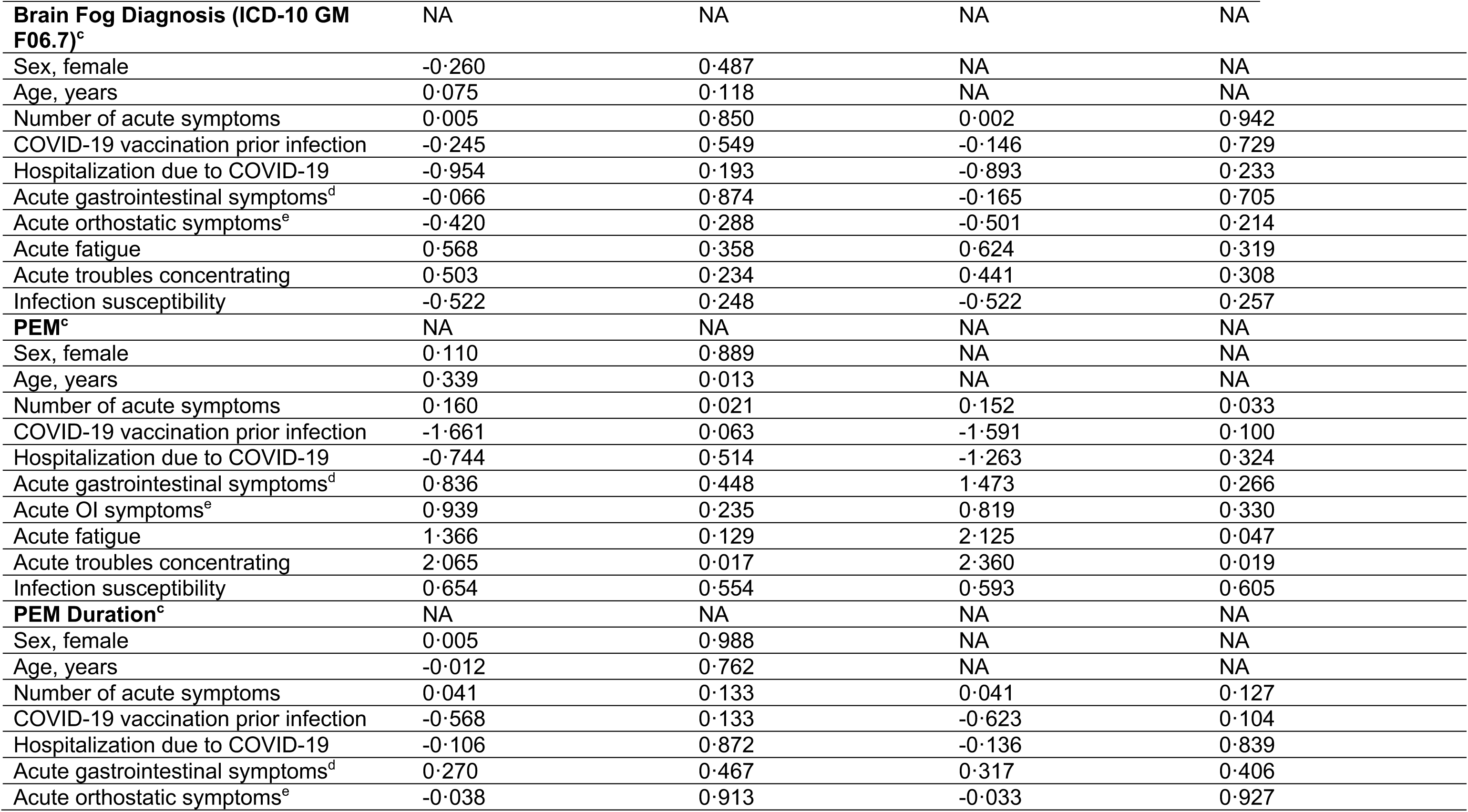
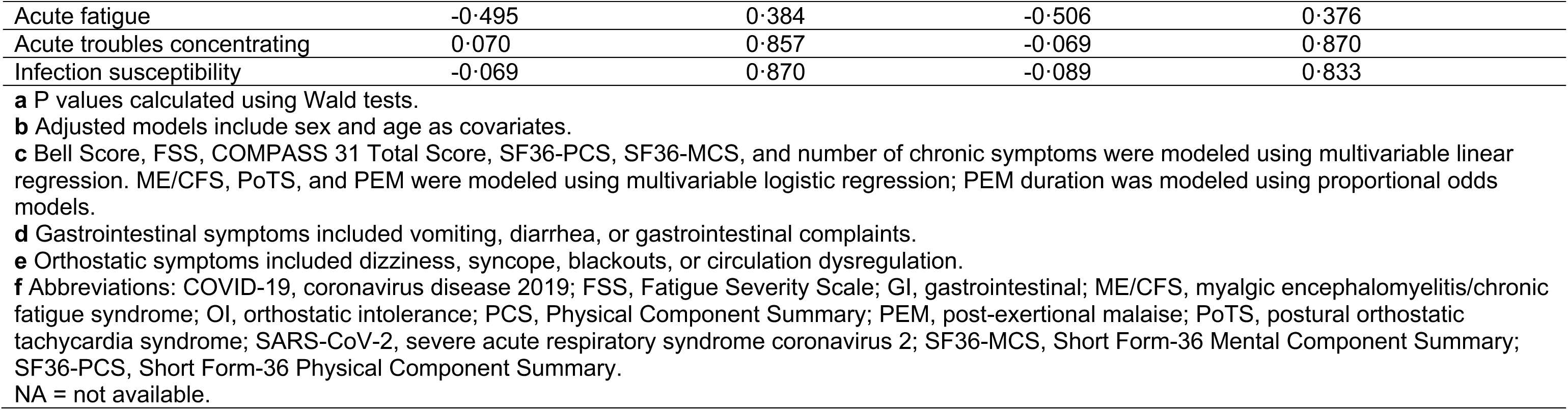
Risk Factor Analyses.

|  | **Unadj. Coefficient /**<br>**Unadj. Log Odds Ratio<sup>b</sup>** | **P-Value<sup>a</sup>** | **Adj. Coefficient /**<br>**Adj. Log Odds Ratio<sup>b</sup>** | **P-Value<sup>a</sup>** |
| --- | --- | --- | --- | --- |
| **Bell Score<sup>c</sup>** | NA | NA | NA | NA |
| Sex, female | 4.238 | 0.202 | NA | NA |
| Age, years | 0.431 | 0.292 | NA | NA |
| Number of acute symptoms | -0.428 | 0.090 | -0.439 | 0.081 |
| COVID-19 vaccination prior infection | -0.537 | 0.882 | 0.266 | 0.942 |
| Hospitalization due to COVID-19 | -7.512 | 0.252 | -8.991 | 0.176 |
| Acute gastrointestinal symptoms<sup>d</sup> | -1.426 | 0.700 | -0.201 | 0.958 |
| Acute orthostatic symptoms<sup>e</sup> | -3.391 | 0.323 | -3.524 | 0.304 |
| Acute fatigue | 7.511 | 0.177 | 7.446 | 0.180 |
| Acute troubles concentrating | -5.595 | 0.133 | -5.067 | 0.176 |
| Infection susceptibility | -3.406 | 0.398 | -3.549 | 0.378 |
| **Fatigue Severity Scale<sup>c</sup>** | NA | NA | NA | NA |
| Sex, female | -0.141 | 0.416 | NA | NA |
| Age, years | 0.004 | 0.862 | NA | NA |
| Number of acute symptoms | 0.045 | <0.001 | 0.045 | <0.001 |
| COVID-19 vaccination prior infection | 0.038 | 0.840 | 0.044 | 0.818 |
| Hospitalization due to COVID-19 | 0.461 | 0.231 | 0.461 | 0.166 |
| Acute gastrointestinal symptoms<sup>d</sup> | 0.232 | 0.232 | 0.196 | 0.331 |
| Acute orthostatic symptoms<sup>e</sup> | 0.383 | 0.037 | 0.377 | 0.042 |
| Acute fatigue | -0.040 | 0.895 | -0.028 | 0.929 |
| Acute troubles concentrating | 0.522 | 0.008 | 0.502 | 0.013 |
| Infection susceptibility | -0.214 | 0.314 | -0.207 | 0.332 |
| **COMPASS 31 Total Score<sup>c</sup>** | NA | NA | NA | NA |
| Sex, female | 8.626 | 0.007 | NA | NA |
| Age, years | 0.016 | 0.969 | NA | NA |
| Number of acute symptoms | 0.620 | 0.014 | 0.633 | 0.010 |
| COVID-19 vaccination prior infection | 1.792 | 0.619 | 1.334 | 0.706 |
| Hospitalization due to COVID-19 | 8.373 | 0.171 | 4.249 | 0.492 |
| Acute gastrointestinal symptoms<sup>d</sup> | -3.279 | 0.375 | -0.773 | 0.838 |
| Acute orthostatic symptoms<sup>e</sup> | 12.346 | <0.001 | 12.570 | <0.001 |
| Acute fatigue | -13.845 | 0.015 | -14.791 | 0.007 |
| Acute troubles concentrating | 4.447 | 0.235 | 5.597 | 0.127 |
| Infection susceptibility | -1.651 | 0.690 | -1.897 | 0.635 |
| **SF36 – PCS<sup>c</sup>** | NA | NA | NA | NA |
| Sex, female | 1.232 | 0.562 | NA | NA |
| Age, years | -0.005 | 0.984 | NA | NA |
| Number of acute symptoms | -0.514 | 0.001 | -0.522 | 0.001 |
| COVID-19 vaccination prior infection | -1.260 | 0.557 | -1.329 | 0.557 |
| Hospitalization due to COVID-19 | -0.669 | 0.853 | -1.111 | 0.764 |
| Acute gastrointestinal symptoms<sup>d</sup> | -3.041 | 0.201 | -2.818 | 0.259 |
| Acute orthostatic symptoms<sup>e</sup> | -3.417 | 0.116 | -3.492 | 0.112 |
| Acute fatigue | -2.021 | 0.595 | -2.115 | 0.581 |
| Acute troubles concentrating | -6.317 | 0.007 | -6.323 | 0.007 |
| Infection susceptibility | -5.304 | 0.028 | -5.335 | 0.029 |
| **SF36 – MCS<sup>c</sup>** | NA | NA | NA | NA |
| Sex, female | -4.469 | 0.054 | NA | NA |
| Age, years | -0.978 | 0.730 | NA | NA |
| Number of acute symptoms | -0.337 | 0.059 | -0.335 | 0.060 |
| COVID-19 vaccination prior infection | 3.234 | 0.185 | 3.446 | 0.170 |
| Hospitalization due to COVID-19 | -8.990 | 0.020 | -7.685 | 0.048 |
| Acute gastrointestinal symptoms<sup>d</sup> | -1.435 | 0.583 | -2.875 | 0.286 |
| Acute orthostatic symptoms<sup>e</sup> | -6.344 | 0.007 | -6.185 | 0.008 |
| Acute fatigue | -4.152 | 0.317 | -3.905 | 0.345 |
| Acute troubles concentrating | -4.699 | 0.070 | -4.788 | 0.063 |
| Infection susceptibility | -1.417 | 0.599 | -1.233 | 0.643 |
| **Number of Chronic Symptoms<sup>c</sup>** | NA | NA | NA | NA |
| Sex, female | 4.074 | 0.089 | NA | NA |
| Age, years | 0.421 | 0.151 | NA | NA |
| Number of acute symptoms | 1.145 | <0.001 | 1.142 | <0.001 |
| COVID-19 vaccination prior infection | -1.963 | 0.449 | -1.303 | 0.618 |
| Hospitalization due to COVID-19 | 6.107 | 0.208 | 4.212 | 0.394 |
| Acute gastrointestinal symptoms<sup>d</sup> | 8.422 | <0.001 | 9.668 | <0.001 |
| Acute orthostatic symptoms<sup>e</sup> | 9.314 | <0.001 | 9.276 | <0.001 |
| Acute fatigue | 4.536 | 0.341 | 3.802 | 0.425 |
| Acute troubles concentrating | 10.151 | <0.001 | 10.900 | <0.001 |
| Infection susceptibility | 4.284 | 0.144 | 4.121 | 0.158 |
| **ME/CFS Diagnosis (ICD-10 GM G93.3)<sup>c</sup>** | NA | NA | NA | NA |
| Sex, female | 1.234 | 0.014 | NA | NA |
| Age, years | 0.068 | 0.206 | NA | NA |
| Number of acute symptoms | 0.010 | 0.765 | 0.010 | 0.762 |
| COVID-19 vaccination prior infection | -0.692 | 0.206 | -0.668 | 0.241 |
| Hospitalization due to COVID-19 | -1.030 | 0.342 | -1.433 | 0.191 |
| Acute gastrointestinal symptoms<sup>d</sup> | 0.065 | 0.893 | 0.371 | 0.476 |
| Acute orthostatic symptoms<sup>e</sup> | -0.423 | 0.341 | -0.444 | 0.334 |
| Acute fatigue | -0.923 | 0.144 | -0.993 | 0.131 |
| Acute troubles concentrating | 0.375 | 0.469 | 0.553 | 0.302 |
| Infection susceptibility | -0.256 | 0.647 | -0.301 | 0.597 |
| **PoTS Diagnosis (ICD-10 GM G90.80)<sup>c</sup>** | NA | NA | NA | NA |
| Sex, female | 0.372 | 0.426 | NA | NA |
| Age, years | -0.055 | 0.355 | NA | NA |
| Number of acute symptoms | 0.068 | 0.071 | 0.074 | 0.051 |
| COVID-19 vaccination prior infection | 0.231 | 0.639 | 0.140 | 0.782 |
| Hospitalization due to COVID-19 | 1.447 | 0.090 | 1.400 | 0.106 |
| Acute gastrointestinal symptoms<sup>d</sup> | 0.047 | 0.926 | 0.195 | 0.718 |
| Acute orthostatic symptoms<sup>e</sup> | 0.249 | 0.606 | 0.353 | 0.476 |
| Acute fatigue | 0.178 | 0.829 | 0.111 | 0.894 |
| Acute troubles concentrating | 0.454 | 0.414 | 0.607 | 0.290 |
| Infection susceptibility | 0.755 | 0.144 | 0.748 | 0.152 |
| **Brain Fog Diagnosis (ICD-10 GM F06.7)<sup>c</sup>** | NA | NA | NA | NA |
| Sex, female | -0.260 | 0.487 | NA | NA |
| Age, years | 0.075 | 0.118 | NA | NA |
| Number of acute symptoms | 0.005 | 0.850 | 0.002 | 0.942 |
| COVID-19 vaccination prior infection | -0.245 | 0.549 | -0.146 | 0.729 |
| Hospitalization due to COVID-19 | -0.954 | 0.193 | -0.893 | 0.233 |
| Acute gastrointestinal symptoms<sup>d</sup> | -0.066 | 0.874 | -0.165 | 0.705 |
| Acute orthostatic symptoms<sup>e</sup> | -0.420 | 0.288 | -0.501 | 0.214 |
| Acute fatigue | 0.568 | 0.358 | 0.624 | 0.319 |
| Acute troubles concentrating | 0.503 | 0.234 | 0.441 | 0.308 |
| Infection susceptibility | -0.522 | 0.248 | -0.522 | 0.257 |
| **PEM<sup>c</sup>** | NA | NA | NA | NA |
| Sex, female | 0.110 | 0.889 | NA | NA |
| Age, years | 0.339 | 0.013 | NA | NA |
| Number of acute symptoms | 0.160 | 0.021 | 0.152 | 0.033 |
| COVID-19 vaccination prior infection | -1.661 | 0.063 | -1.591 | 0.100 |
| Hospitalization due to COVID-19 | -0.744 | 0.514 | -1.263 | 0.324 |
| Acute gastrointestinal symptoms<sup>d</sup> | 0.836 | 0.448 | 1.473 | 0.266 |
| Acute OI symptoms<sup>e</sup> | 0.939 | 0.235 | 0.819 | 0.330 |
| Acute fatigue | 1.366 | 0.129 | 2.125 | 0.047 |
| Acute troubles concentrating | 2.065 | 0.017 | 2.360 | 0.019 |
| Infection susceptibility | 0.654 | 0.554 | 0.593 | 0.605 |
| **PEM Duration<sup>c</sup>** | NA | NA | NA | NA |
| Sex, female | 0.005 | 0.988 | NA | NA |
| Age, years | -0.012 | 0.762 | NA | NA |
| Number of acute symptoms | 0.041 | 0.133 | 0.041 | 0.127 |
| COVID-19 vaccination prior infection | -0.568 | 0.133 | -0.623 | 0.104 |
| Hospitalization due to COVID-19 | -0.106 | 0.872 | -0.136 | 0.839 |
| Acute gastrointestinal symptoms<sup>d</sup> | 0.270 | 0.467 | 0.317 | 0.406 |
| Acute orthostatic symptoms<sup>e</sup> | -0.038 | 0.913 | -0.033 | 0.927 |
| Acute fatigue | -0.495 | 0.384 | -0.506 | 0.376 |
| Acute troubles concentrating | 0.070 | 0.857 | -0.069 | 0.870 |
| Infection susceptibility | -0.069 | 0.870 | -0.089 | 0.833 |

**a** P values calculated using Wald tests.
**b** Adjusted models include sex and age as covariates.
**c** Bell Score, FSS, COMPASS 31 Total Score, SF36-PCS, SF36-MCS, and number of chronic symptoms were modeled using multivariable linear regression. ME/CFS, PoTS, and PEM were modeled using multivariable logistic regression; PEM duration was modeled using proportional odds models.
**d** Gastrointestinal symptoms included vomiting, diarrhea, or gastrointestinal complaints.
**e** Orthostatic symptoms included dizziness, syncope, blackouts, or circulation dysregulation.
**f** Abbreviations: COVID-19, coronavirus disease 2019; FSS, Fatigue Severity Scale; GI, gastrointestinal; ME/CFS, myalgic encephalomyelitis/chronic fatigue syndrome; OI, orthostatic intolerance; PCS, Physical Component Summary; PEM, post-exertional malaise; PoTS, postural orthostatic tachycardia syndrome; SARS-CoV-2, severe acute respiratory syndrome coronavirus 2; SF36-MCS, Short Form-36 Mental Component Summary; SF36-PCS, Short Form-36 Physical Component Summary.
NA = not available.

### Risk Factors Analysis for Severe PCC

The result of the k-means clustering is visualized in **Suppl. Figure S1A (Appendix p.6)**. The first cluster encompassed all 29/70 (41%) patients with confirmed or probable ME/CFS and was further characterized by a lower Bell Score, more severe fatigue (FSS), more autonomic symptoms (COMPASS 31), more PCC symptoms, a lower SF36-PCS and a lower SF36-MCS compared to the second cluster (**Suppl. Figure S1B-S1H**). Female sex, but not age, had a significantly greater likelihood of being attributed to the severe PCC cluster (OR = 3·31, P = 0·031). Adjusted for sex and age, the number of acute symptoms (adj. OR = 1·22, P < 0·001), acute symptoms of OI (adj. OR = 9·87, P = 0·002), and acute troubles concentrating (adj. OR = 11·8, P = 0·005) were significantly associated with the severe PCC cluster. (**Figure 1**)

**Figure 1.**
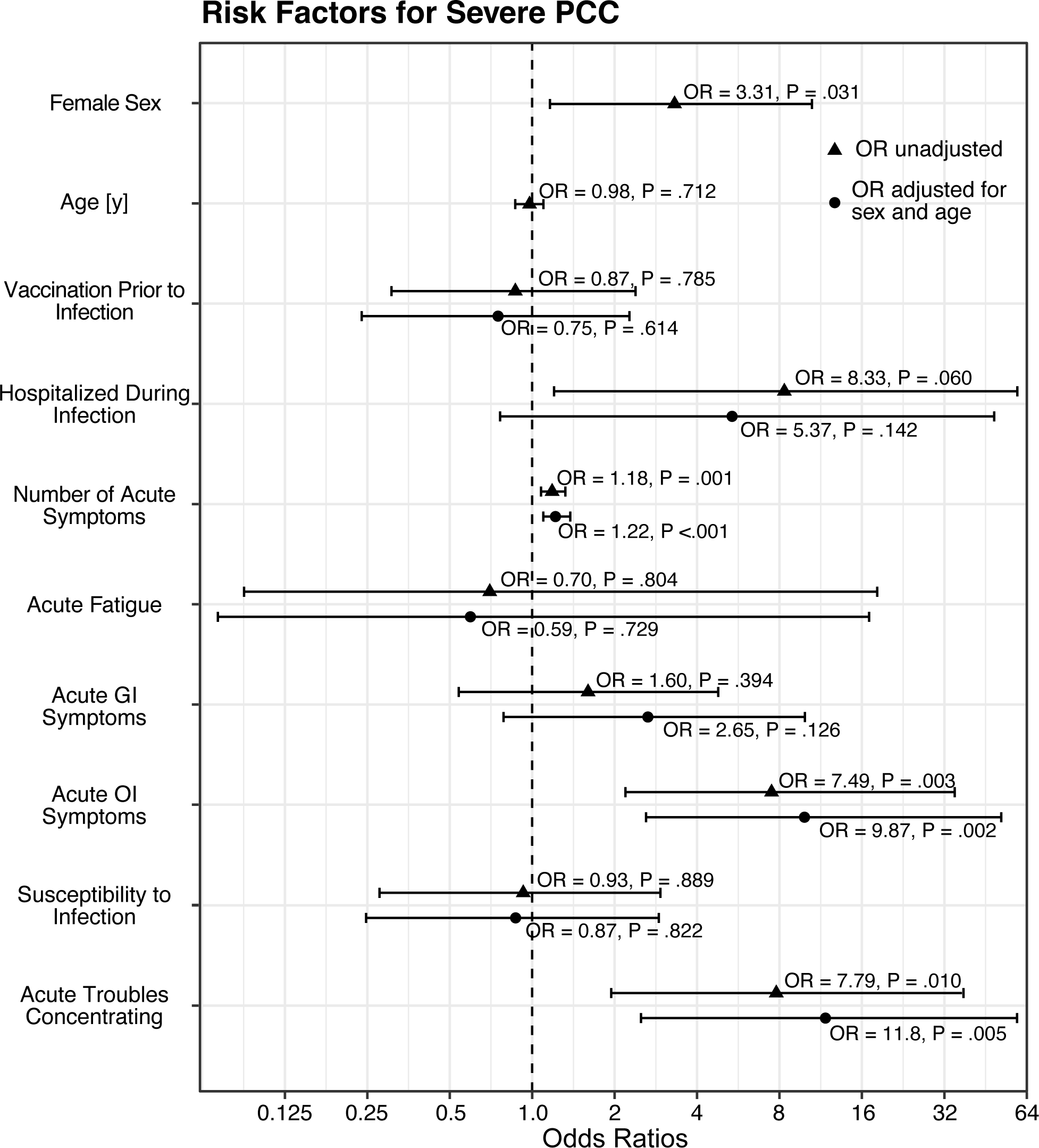
Forest plot of predictors for severe vs. moderate PCC_cyp_ (N=70) derived from logistic regression models. Circles show unadjusted odds ratios (OR), triangles show OR adjusted for sex and age. Bars indicate 95%-confidence intervals.

A trained and cross-validated LASSO retained the following predictors a final logistic regression model: female sex (coefficient: 1·08), age (coefficient: -0·07), hospitalization during acute COVID-19 (coefficient: 0·10), number of acute COVID-19 symptoms (coefficient: 0·10), acute OI symptoms (coefficient: 1·09), acute fatigue (coefficient: -0·95), and acute troubles concentrating (coefficient: 0·80). A ROC analysis of this model resulted in an AUC of 85·7%. The optimal threshold for predicting severe PCC was if the predicted probability p ≥63·8% (Youden index; **Suppl. Figure S1I**). The model reliably detected CYP with “severe PCC” (sensitivity: 65·6%) or “moderate PCC” (specificity: 90·2%), yielding an overall accuracy of 80·0% (balanced accuracy: 77·9%) (**Suppl. Figure S1J, S1K**). A binary categorization of the acute number of symptoms in the penalized logistic regression model revealed that ≥12 acute COVID-19 symptoms had the best model fit, indicated by minimal cross-validated model error (**Suppl. Figure S1L**).

## DISCUSSION

This study identified key risk factors for severe PCC in CYP after confirmed or probable COVID-19, highlighted a distinct subgroup of highly affected individuals, and introduced the novel MLCSQ as a robust instrument for standardized assessment of PCC symptoms. The most important finding was that a high number of acute symptoms and the presence of autonomic symptoms during the acute phase of initial SARS-CoV-2 infection were early and specific predictors of severe PCC and thereby may allow for tailored and timely symptom-oriented care.

A higher number of acute initial symptoms was significantly associated with more severe long-term outcomes, including more severe fatigue (FSS) and autonomic dysfunction (COMPASS-31), reduced physical and mental HRQoL (SF-36), and a higher number of chronic symptoms (MLCSQ). These results expand on earlier work, which indicated that acute EBV-IM symptom complexity and autonomic EBV-IM symptoms predicted prolonged recovery and/or chronic fatigue.^23^ The latter finding was consistent with results from Jason et al., who identified greater symptom burden and immune activation as predictors of EBV-IM-driven ME/CFS.

In adults, the presence of five acute COVID-19 symptoms has been associated with increased PCC risk.^18^ Moreover, symptomatic versus asymptomatic SARS-CoV-2 infection has been linked to long-term sequelae in adults and children. While earlier pediatric studies identified hospitalization for COVID-19^3,19,20^ and acute complications such as pneumonia, reduced oxygen saturation, and hypotension as risk factors for PCC, our data are the first to demonstrate that a threshold of ≥12 COVID-19 symptoms allows for identifying CYP at risk of severe PCC.

Among acute COVID-19 symptoms, symptoms of OI (dizziness, syncope, blackouts, and circulatory dysregulation) emerged as another strong predictor of severe PCC. These symptoms were significantly associated with increased fatigue, greater autonomic distress, poorer mental health, and a higher number of symptoms in the context of PCC. This supports the hypothesis that autonomic dysregulation plays a key role in the development of post-viral sequelae.^14^

Unsupervised clustering delineated a clinically coherent “severe” phenotype that concentrated all ME/CFS cases and showed markedly worse fatigue, autonomic symptom burden, functional status, and HRQoL than the comparison cluster. This supports the robustness of our cluster endpoint.

The risk-factor analyses found that female sex was associated with higher odds of severe PCC, which is consistent with prior pediatric reports.^8^ In contrast to several pediatric cohort studies reporting, we could not confirm age as a relevant risk factor for PCC. ^3^ E.g., Rao et al. found that children younger than five years were more strongly associated with PASC.^19^ This controversy might indicate that more nuanced age-specific modelling is needed. Acute orthostatic-intolerance symptoms and acute troubles concentrating exhibited the strongest adjusted associations with severe PCC, highlighting early autonomic and neurocognitive involvement as salient signals. A higher acute symptom burden also tracked with risk, with the penalized model indicating ≥12 acute symptoms as a pragmatic cut-point. It is not surprising that acute fatigue was not identified as a discriminatory risk factor since it was prevalent in 90% of all patients.

LASSO-penalized logistic regression selected seven independent predictors for severe PCC, including female sex, younger age, hospitalization, number of acute symptoms, acute OI symptoms, acute troubles concentrating, and absence of acute fatigue. In accordance, female sex has been considered a risk factor in pediatric patients.^8^ Interestingly, acute fatigue and sex now appeared as risk factors. However, they may merely reflect an additional effect in the presence of the remaining information of the other predictors in the model. This model surely needs further evaluation in further studies.

All ME/CFS patients included in the cluster analysis were assigned to the severe PCC cluster. This subgroup of ME/CFS patients was characterized by older age, female predominance, higher acute symptom burden, and more severe scores across all domains. This aligns with previous studies reporting that ME/CFS is often associated with OI in adolescents,^10^ and that pediatric PCC patients with ME/CFS, compared to non-ME/CFS, had a higher symptom load.^5^

### Strengths and Limitations

A key strength of our work is its clinical immediacy: we identify a pragmatic threshold, i.e., ≥12 acute symptoms, that enables early risk stratification for severe PCC in CYP. The study leverages a relatively large, well-characterized pediatric cohort and a comprehensive, multidimensional assessment using validated clinical instruments and PROMs, enhancing reproducibility and comparability. Our risk model is robust and data-driven, combining unsupervised clustering with LASSO and cross-validation to prioritize predictors with joint explanatory value. Finally, the retained predictors (sex, age, hospitalization, acute symptom load, acute OI symptoms, acute concentration problems, and acute fatigue) are easy to assess, supporting direct clinical translation.

Unfortunately, this analysis did not include a control group that fully recovered after SARS-CoV-2 infection. Therefore, risk factors were conditional on PCC development, and general extrapolation to CYP with SARS-CoV-2 infections remains limited. Moreover, COVID-19 symptoms might be influenced by recall bias, and the proposed clinical cut-off of ≥12 acute symptoms requires further validation, including age- and sex-specific analyses in larger, independent cohorts. Additional limitations include missing data in some outcome variables, which reduced the effective sample size for clustering and regression analyses and may have affected the stability of model estimates. Lastly, findings for our German monocenter cohort may not be generalizable to other populations or healthcare systems.

This study highlights the importance of acute-phase symptom profiles, particularly the number of symptoms and autonomic dysfunction, for identifying CYP at risk for severe PCC. The novel MLCSQ enables a standardized, quantitative assessment of PCC symptom load. Our findings emphasize the need for systematic early symptom documentation to guide timely risk stratification, individualized follow-up, and targeted intervention in pediatric populations affected by long-term sequelae of COVID-19.

## Data Availability

All data supporting the findings of this study is reported in this published manuscript and its supplementary Appendix. Ethics approval and patient consent forms do not allow individual participant data to be made publicly available. Requests for further information can be directed to the corresponding author.

## Contributors

Q.D., Ma.Hä., D.S., C.C., M.A., LC.P., S.S., K.G., R.P., L.M., and U.B. conceptualized the study and developed the methodology. Data acquisition and curation were performed by Q.D., Ma.Hä., D.S., T.W., C.C., A.Gr., A.L., C.I., Ca.W., M.E., S.B., N.B., H.W., H.H., LC.P., L.B., S.S., Co.Wa., M.A., Ar.Gr., R.P., and U.B.. Formal analyses were conducted by Q.D., L.M., and U.B.. All authors contributed to data validation of the study. Supervision was provided by U.B. and L.M., with project administration and funding acquisition by U.B. Software and visualisation were handled by Q.D. and L.M.. The original draft was written by Q.D., L.M., and U.B., and all authors reviewed and edited the manuscript critically. U.B., L.M., and Q.D. have directly accessed and verified all underlying data. All authors had full access to all study data, accept responsibility for the decision to submit for publication, and agree to be accountable for all aspects of the work.

