## Appendix for "Risk Factors for Severe Post-COVID Condition in Children, Adolescents, and Young Adults"

**Contents**

**Suppl. Table S1.** *Post-COVID-Associated Diagnoses (ICD-10-GM U09.9!)*

**Suppl. Table S2.** *Comparison of PCC Patients with and without ME/CFS*

**Suppl. Figure S1.** *Clustering and prediction modelling of severe PCC outcomes*

**Suppl. Material M1.** *Munich Long-COVID Symptom Questionnaire (MLCSQ)*

**Supplementary Table S1.** *Post-COVID-associated ICD-10 GM Diagnoses (U09.9!)*

| | Overall[a]<br><br>N = 120 | Children[a]<br>Age 7 to 11 years<br>N = 16 | Adolescents[a]<br>Age 12 to 17 years<br>N = 71 | Adults[a]<br>Age 18 to 25 years<br>N = 33 | P-Value[b] |
|---|---|---|---|---|---|
| R53 – Fatigue | 81 / 120 (68%) | 12 / 16 (75%) | 51 / 71 (72%) | 18 / 33 (55%) | 0·170 |
| F06 – Brain fog (F06.7) | 64 / 120 (53%) | 10 / 16 (63%) | 31 / 71 (44%) | 23 / 33 (70%) | 0·034 |
| R51 – Headache | 49 / 120 (41%) | 10 / 16 (63%) | 26 / 71 (37%) | 13 / 33 (39%) | 0·160 |
| R42 – Vertigo | 32 / 120 (27%) | 4 / 16 (25%) | 21 / 71 (30%) | 7 / 33 (21%) | 0·731 |
| R06 – Dyspnoea (R06.0) | 25 / 120 (21%) | 3 / 16 (19%) | 16 / 71 (23%) | 6 / 33 (18%) | 0·948 |
| G47 – Sleep disorders | 20 / 120 (17%) | 4 / 16 (25%) | 10 / 71 (14%) | 6 / 33 (18%) | 0·502 |
| M79 – Myalgia (M79.1) | 17 / 120 (14%) | 4 / 16 (25%) | 7 / 71 (10%) | 6 / 33 (18%) | 0·199 |
| F43 – Reaction to severe stress, and adjustment disorders | 15 / 120 (13%) | 1 / 16 (6%) | 6 / 71 (9%) | 8 / 33 (24%) | 0·091 |
| R10 – Abdominal pain (R10.4) | 9 / 120 (8%) | 2 / 16 (13%) | 5 / 71 (7%) | 2 / 33 (6%) | 0·776 |
| R11 – Nausea and vomiting | 9 / 120 (8%) | 4 / 16 (25%) | 5 / 71 (7%) | 0 / 33 (0%) | 0·008 |
| R43 – Disturbances of smell and taste (R43.8) | 9 / 120 (8%) | 1 / 16 (6%) | 4 / 71 (6%) | 4 / 33 (12%) | 0·488 |
| G47 – Hypersomnia (G47.1) | 8 / 120 (7%) | 1 / 16 (6%) | 5 / 71 (7%) | 2 / 33 (6%) | >0·999 |
| M79 – Pain in limb (M79.6) | 7 / 120 (6%) | 4 / 16 (25%) | 2 / 71 (3%) | 1 / 33 (3%) | 0·009 |
| M25 – Pain in joint (M25.5) | 6 / 120 (5%) | 3 / 16 (19%) | 2 / 71 (3%) | 1 / 33 (3%) | 0·038 |
| R00 – Palpitations (R00.2) | 5 / 120 (4%) | 1 / 16 (6%) | 2 / 71 (3%) | 2 / 33 (6%) | 0·515 |

**a** Data are presented as n/N (%).Percentages may not total 100% because of rounding.
**b** P-values were calculated using the Fisher exact test, or Pearson $\chi^2$ test, as appropriate.
**c** Diagnoses are reported only if they occurred in ≥5 patients.
**d** Abbreviations: ICD-10, International Statistical Classification of Diseases and Related Health Problems, 10th Revision; PCC, pediatric post–COVID-19 condition.

NA = not available.

**Supplementary Table S2.** *Comparison of PCC Patients with and without ME/CFS*

| | MECFS[a] (N = 29) | non-MECFS[a] (N = 91) | P-Value[b] |
|---|---|---|---|
| Sex (female) | 23/29 (79%) | 48/91 (53%) | 0·016 |
| Age | 15·0 (14·0 - 19·0) | 15·0 (13·0 - 17·0) | 0·224 |
| Number of 83 possible PCC symptoms reported | 33 (25·3 - 39·3) | 29 (19·5 - 36) | 0·065 |
| Participation | NA | NA | 0·003 |
| full participation | 0 / 28 (0%) | 17 / 91 (19%) | NA |
| > 50% participation | 4 / 28 (14%) | 29 / 91 (32%) | NA |
| < 50% participation | 12 / 28 (43%) | 28 / 91 (31%) | NA |
| no participation | 12 / 28 (43%) | 17 / 91 (19%) | NA |
| Bell-Score | 30·0 (30·0 - 50·0) | 50·0 (40·0 - 60·0) | <0·001 |
| Fatigue Severity Scale | 6·6 (6·3 - 6·8) | 6·2 (5·7 - 6·6) | 0·012 |
| PEM screening positive | 29 / 29 (100%) | 83 / 91 (91%) | 0·274 |
| PEM Duration (hours) | NA | NA | 0·001 |
| < 1 | 0 / 28 (0%) | 5 / 86 (6%) | NA |
| 2 - 3 | 2 / 28 (7%) | 29 / 86 (34%) | NA |
| 4 - 10 | 4 / 28 (14%) | 24 / 86 (28%) | NA |
| 11 - 13 | 2 / 28 (7%) | 6 / 86 (7%) | NA |
| 14 - 23 | 9 / 28 (31%) | 10 / 86 (12%) | NA |

| | | | |
|---|---|---|---|
| ≥ 24 | 11 / 28 (38%) | 12 / 86 (14%) | NA |
| COMPASS 31 Total score | 34·9 (25·6 - 43·5) | 25·6 (16·8 - 36·5) | 0·032 |
| SF36-PCS | 27·1 (23·6 - 32·0) | 31·5 (28·3 - 38·6) | 0·025 |
| SF36-MCS | 42·4 (29·5 - 47·9) | 45·6 (36·0 - 52·7) | 0·114 |
| PoTS (ICD-10 GM G90.80) | 9 / 29 (31%) | 16 / 91 (18%) | 0·278 |

**a** Data are expressed as median (IQR) or n/N (%), as appropriate. Percentages may not total 100% because of rounding.
**b** P-values were calculated using the Kruskal-Wallis rank sum test, Fisher exact test, or Pearson $\chi^2$ test, as appropriate.
**c** Abbreviations: ME/CFS, myalgic encephalomyelitis/chronic fatigue syndrome; PCC, pediatric post–COVID-19 condition; FSS, Fatigue Severity Scale; PEM, post-exertional malaise; SF36-PCS, SF-36 Physical Component Summary; SF36-MCS, SF-36 Mental Component Summary; PoTS, postural orthostatic tachycardia syndrome.
NA = not available.

**Suppl. Figure S1.** *Clustering and prediction modelling of severe PCC outcomes*

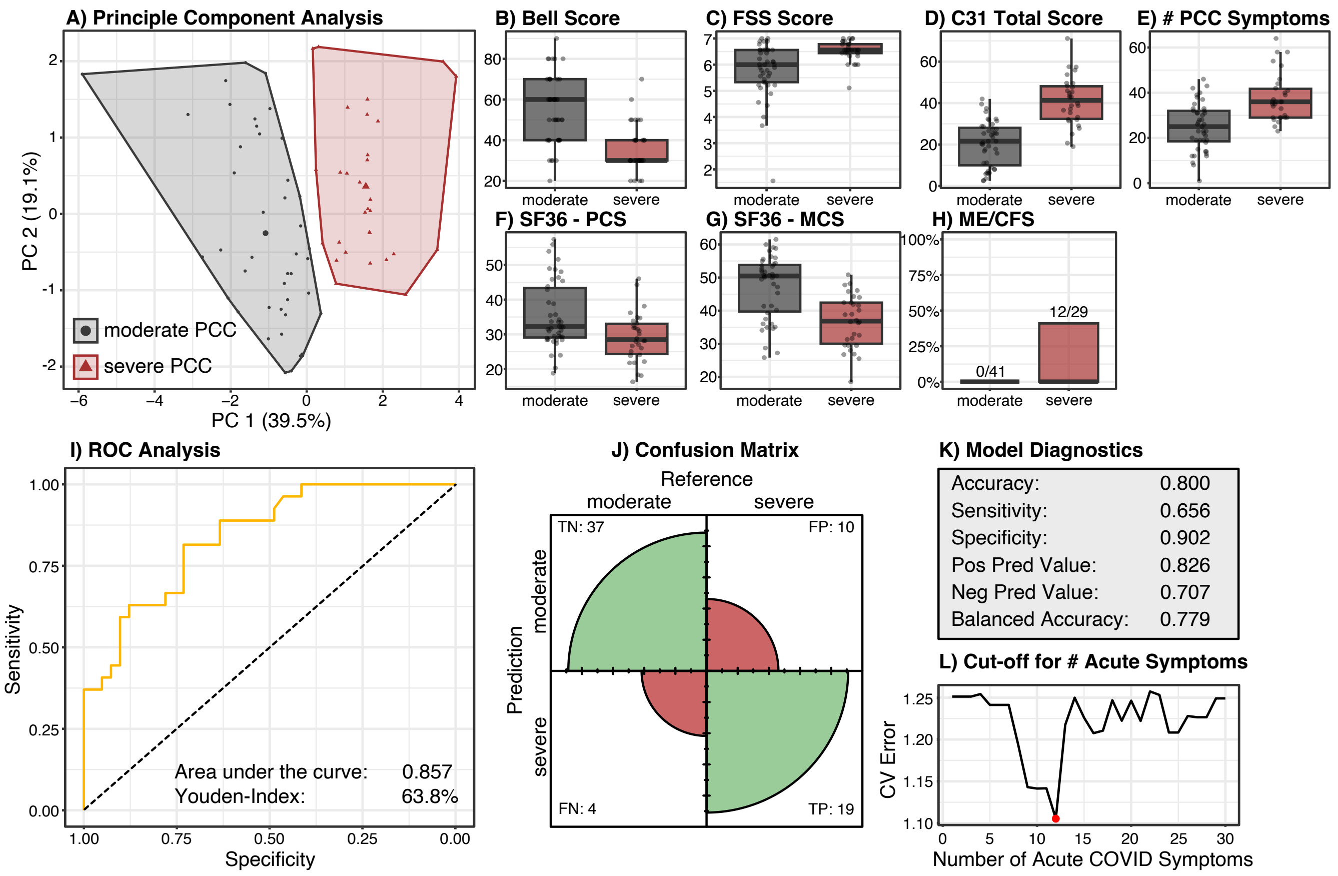

*(A) Principle component analysis showing the result of the k-means clustering of the (scaled) outcome variables (Bell Score, Fatigue Severity Scale (FSS), COMPASS 31 (C31) Total Score, Number (#) of PCC symptoms, SF36-PCS, SF36-MSS, ME/CFS diagnosis). (B) – (H) comparisons of outcome variables between patient clusters with "moderate PCC" and "severe PCC". (I) ROC analysis, (J) confusion matrix, and (K) model diagnostics of the resulting LASSO-penalized logistic regression model. (L) Estimation of the optimal cut-off values for the acute number of COVID-19 symptoms by determining the minimal leave-one-out cross-validation (CV) error (y-axis) of LASSO-penalized logistic regression models with the respective cut-off number (x-axis). The red dot marks the minimal CV error at a number of ≥12 acute COVID-19 symptoms.*

Klinikum rechts der Isar Technical University of Munich

### Munich Long COVID Symptom Questionnaire (MLCSQ)

| | |
|---|---|
| Name: | Name (Physician): |
| First Name: | First Name (Physician): |
| Date of Birth: | Date (Physician): |
| Date of Completion: Completion Time: min | Institution: |

***Please complete the questionnaire as much as possible by yourself and ask your parents for help if needed. Any remaining open questions or issues of understanding should be clarified during the medical consultation.***

| | | Did not occur since the SARS-CoV-2 infection | If symptoms have occurred since the SARS-CoV-2 infection, please answer the following questions: How frequently does this symptom occur? | How severe is the symptom? 1 = mild 2 = moderate 3 = severe | Medical notes |
|---|---|---|---|---|---|
| **I** | **Fatigue / Exercise intolerance** | | | | |
| **1** | Fatigue (exhaustion)[1,2] | 0 | □ Not anymore □ Intermittent □ Persistent | 1 2 3 | |
| **2** | Worsening of symptoms following mild mental and/or physical activity (exercise intolerance)[1] | 0 | □ Not anymore □ Intermittent □ Persistent | 1 2 3 | |
| **3** | Reduced endurance[2] | 0 | □ Not anymore □ Intermittent □ Persistent | 1 2 3 | |
| **II** | **Ears-Nose-Throat (ENT)** | | | | |
| **4** | Sore throat[2] | 0 | □ Not anymore □ Intermittent □ Persistent | 1 2 3 | |
| **5** | Swollen lymph nodes | 0 | □ Not anymore □ Intermittent □ Persistent | 1 2 3 | |
| **6** | Voice disorder | 0 | □ Not anymore □ Intermittent □ Persistent | 1 2 3 | |
| **7** | Problem swallowing[1] | 0 | □ Not anymore □ Intermittent □ Persistent | 1 2 3 | |
| **8** | Dryness of the mouth[3] | 0 | □ Not anymore □ Intermittent □ Persistent | 1 2 3 | |
| **9** | Cold[2] | 0 | □ Not anymore □ Intermittent □ Persistent | 1 2 3 | |
| **10** | Sinus disorder[2] | 0 | □ Not anymore □ Intermittent □ Persistent | 1 2 3 | |
| **11** | Loss of smell[1,2] | 0 | □ Not anymore □ Intermittent □ Persistent | 1 2 3 | |
| **12** | Loss of taste[1,2] | 0 | □ Not anymore □ Intermittent □ Persistent | 1 2 3 | |
| **13** | Altered smell[1,2] | 0 | □ Not anymore □ Intermittent □ Persistent | 1 2 3 | |
| **14** | Altered taste[1,2] | 0 | □ Not anymore □ Intermittent □ Persistent | 1 2 3 | |
| **15** | Ear pain[2] | 0 | □ Not anymore □ Intermittent □ Persistent | 1 2 3 | |
| **16** | Hearing impairment[1] | 0 | □ Not anymore □ Intermittent □ Persistent | 1 2 3 | |
| **17** | Ringing in ears[1,2] | 0 | □ Not anymore □ Intermittent □ Persistent | 1 2 3 | |
| **18** | Further ENT complaints: | | | | |
| **III** | **Ophthalmic** | | | | |
| **19** | Conjunctivitis | 0 | □ Not anymore □ Intermittent □ Persistent | 1 2 3 | |
| **20** | Dry eyes[3] | 0 | □ Not anymore □ Intermittent □ Persistent | 1 2 3 | |
| **21** | Light sensitivity[3] | 0 | □ Not anymore □ Intermittent □ Persistent | 1 2 3 | |
| **22** | Vision impairment[1,3] | 0 | □ Not anymore □ Intermittent □ Persistent | 1 2 3 | |
| **23** | Further ophthalmic complaints: | | | | |
| **IV** | **Pulmonary** | | | | |
| **24** | Persistent dry cough[1,2] | 0 | □ Not anymore □ Intermittent □ Persistent | 1 2 3 | |
| **25** | Congestion | 0 | □ Not anymore □ Intermittent □ Persistent | 1 2 3 | |
| **26** | Pain on breathing[1] | 0 | □ Not anymore □ Intermittent □ Persistent | 1 2 3 | |
| **27** | Shortness of breath at rest[1,2] | 0 | □ Not anymore □ Intermittent □ Persistent | 1 2 3 | |
| **28** | Shortness of breath with activity[1,2] | 0 | □ Not anymore □ Intermittent □ Persistent | 1 2 3 | |
| **29** | Further pulmonary complaints: | | | | |

| | | Did not occur since the SARS-CoV-2 infection | If symptoms have occurred since the SARS-CoV-2 infection, please answer the following questions: How frequently does this symptom occur? | How severe is the symptom? 1 = mild 2 = moderate 3 = severe | Medical notes |
|---|---|---|---|---|---|
| **V** | **Cardiovascular** | | | | |
| **30** | Chest tightness[2] | 0 | □ Not anymore □ Intermittent □ Persistent | 1 2 3 | |
| **31** | Breath-independent chest pain[1,2] | 0 | □ Not anymore □ Intermittent □ Persistent | 1 2 3 | |
| **32** | Heart pounding[1,2] | 0 | □ Not anymore □ Intermittent □ Persistent | 1 2 3 | |
| **33** | Heart racing[1,2] | 0 | □ Not anymore □ Intermittent □ Persistent | 1 2 3 | |
| **34** | Orthostatic intolerance[3] | 0 | □ Not anymore □ Intermittent □ Persistent | 1 2 3 | |
| **35** | Fainting and/or Blackouts[1] | 0 | □ Not anymore □ Intermittent □ Persistent | 1 2 3 | |
| **36** | Further cardiovascular complaints: | | | | |
| **VI** | **Gastro-intestinal** | | | | |
| **37** | Stomach pain[1,2,3] | 0 | □ Not anymore □ Intermittent □ Persistent | 1 2 3 | |
| **38** | Nausea[1,2] | 0 | □ Not anymore □ Intermittent □ Persistent | 1 2 3 | |
| **39** | Vomiting[1,2] | 0 | □ Not anymore □ Intermittent □ Persistent | 1 2 3 | |
| **40** | Diarrhoea[1,2,3] | 0 | □ Not anymore □ Intermittent □ Persistent | 1 2 3 | |
| **41** | Constipation[1,3] | 0 | □ Not anymore □ Intermittent □ Persistent | 1 2 3 | |
| **42** | Bloatedness[3] | 0 | □ Not anymore □ Intermittent □ Persistent | 1 2 3 | |
| **43** | Flatulence[3] | 0 | □ Not anymore □ Intermittent □ Persistent | 1 2 3 | |
| **44** | Loss of appetite[1,2] | 0 | □ Not anymore □ Intermittent □ Persistent | 1 2 3 | |
| **45** | Weight loss[1,2] | 0 | □ Not anymore □ Intermittent □ Persistent | 1 2 3 | |
| **46** | Further gastro-intestinal complaints: | | | | |
| **VII** | **Musculoskeletal** | | | | |
| **47** | Muscle pain[1,2] | 0 | □ Not anymore □ Intermittent □ Persistent | 1 2 3 | |
| **48** | Muscle weakness[1] | 0 | □ Not anymore □ Intermittent □ Persistent | 1 2 3 | |
| **49** | Muscle twitching[1] | 0 | □ Not anymore □ Intermittent □ Persistent | 1 2 3 | |
| **50** | Muscle tremor[1] | 0 | □ Not anymore □ Intermittent □ Persistent | 1 2 3 | |
| **51** | Muscle stiffness[1] | 0 | □ Not anymore □ Intermittent □ Persistent | 1 2 3 | |
| **52** | Joint pain[1,2] | 0 | □ Not anymore □ Intermittent □ Persistent | 1 2 3 | |
| **53** | Joint swelling[1,2] | 0 | □ Not anymore □ Intermittent □ Persistent | 1 2 3 | |
| **54** | Redness and/or overheating of joints[2] | 0 | □ Not anymore □ Intermittent □ Persistent | 1 2 3 | |
| **55** | Back pain | 0 | □ Not anymore □ Intermittent □ Persistent | 1 2 3 | |
| **56** | Further musculoskeletal complaints: | | | | |
| **VIII** | **Neurological** | | | | |
| **57** | Headaches[1,2] | 0 | □ Not anymore □ Intermittent □ Persistent | 1 2 3 | |
| **58** | Trouble in concentrating[1,2] | 0 | □ Not anymore □ Intermittent □ Persistent | 1 2 3 | |
| **59** | Forgetfulness[1,2] | 0 | □ Not anymore □ Intermittent □ Persistent | 1 2 3 | |
| **60** | Light-headedness[1] | 0 | □ Not anymore □ Intermittent □ Persistent | 1 2 3 | |
| **61** | Dizziness[1,2] | 0 | □ Not anymore □ Intermittent □ Persistent | 1 2 3 | |
| **62** | Numbness or tingling[1,2] | 0 | □ Not anymore □ Intermittent □ Persistent | 1 2 3 | |
| **63** | Can't feel one side of body or face[1] | 0 | □ Not anymore □ Intermittent □ Persistent | 1 2 3 | |
| **64** | Can't move one side of body or face [1] | 0 | □ Not anymore □ Intermittent □ Persistent | 1 2 3 | |
| **65** | Slowness of movement[1] | 0 | □ Not anymore □ Intermittent □ Persistent | 1 2 3 | |
| **66** | Problems with balance[1] | 0 | □ Not anymore □ Intermittent □ Persistent | 1 2 3 | |
| **67** | Problems with gait/falls[1] | 0 | □ Not anymore □ Intermittent □ Persistent | 1 2 3 | |
| **68** | Seizures[1] | 0 | □ Not anymore □ Intermittent □ Persistent | 1 2 3 | |
| **69** | Further neurological complaints: | | | | |
| **IX** | **Body Temperature** | | | | |
| **70** | Increased sweating[2,3] | 0 | □ Not anymore □ Intermittent □ Persistent | 1 2 3 | |
| **71** | Fever > 38,0 °C[1,2] | 0 | □ Not anymore □ Intermittent □ Persistent | 1 2 3 | |
| **72** | Chills | 0 | □ Not anymore □ Intermittent □ Persistent | 1 2 3 | |
| **73** | Further body temperature complaints: | | | | |

| | Did not occur since the SARS-CoV-2 infection | If symptoms have occurred since the SARS-CoV-2 infection, please answer the following questions<br>How frequently does this symptom occur? | How severe is the symptom?<br>1 = mild<br>2 = moderate<br>3 = severe | Medical notes |
|---|---|---|---|---|
| **X Dermatological** | | | | |
| **74** Skin rash[1,2] | 0 | □ Not anymore □ Intermittent □ Persistent<br>Affected areas: □ Face □ Buttocks □ Legs □ Toes □ Trunk □ Arms □ Fingers | 1 2 3 | |
| **75** COVID toes (purple, pink, bluish)[1,2,3] | 0 | □ Not anymore □ Intermittent □ Persistent | 1 2 3 | |
| **76** Other changes in skin color on hands and/or feet (red, white)[1,3] | 0 | □ Not anymore □ Intermittent □ Persistent | 1 2 3 | |
| **77** Swollen ankles[1] / Oedemas | 0 | □ Not anymore □ Intermittent □ Persistent | 1 2 3 | |
| **78** Hair loss[2] | 0 | □ Not anymore □ Intermittent □ Persistent | 1 2 3 | |
| **79** Further dermatological complaints: | | | | |

| | | | |
|---|---|---|---|
| **XI Sleep** | | | |
| **80** Sleeping more[1,2] | 0 | □ Not anymore □ Intermittent □ Persistent | 1 2 3 |
| **81** Sleeping less[1,2] | 0 | □ Not anymore □ Intermittent □ Persistent | 1 2 3 |
| **82** Other sleep disturbances[2] | 0 | □ Not anymore □ Intermittent □ Persistent | 1 2 3 |
| **83** Further sleep complaints: | | | |

| | | | |
|---|---|---|---|
| **XII Mental health** | | | |
| **84** Behaviour change[1] | 0 | □ Not anymore □ Intermittent □ Persistent | 1 2 3 |
| **85** Tic | 0 | □ Not anymore □ Intermittent □ Persistent | 1 2 3 |
| **86** Depressed mood[1,2] | 0 | □ Not anymore □ Intermittent □ Persistent | 1 2 3 |
| **87** Loss of interest/pleasure[1] | 0 | □ Not anymore □ Intermittent □ Persistent | 1 2 3 |
| **88** Listlessness / Lack of motivation[2] | 0 | □ Not anymore □ Intermittent □ Persistent | 1 2 3 |
| **89** Anxiety[1,2] | 0 | □ Not anymore □ Intermittent □ Persistent | 1 2 3 |
| **90** Hallucinations[1] | 0 | □ Not anymore □ Intermittent □ Persistent | 1 2 3 |
| **91** Further mental health complaints: | | | |

| | | | |
|---|---|---|---|
| **XIII Urogenital** | | | |
| **92** Problems passing urine[1,3] | 0 | □ Not anymore □ Intermittent □ Persistent | 1 2 3 |
| To be answered only by patients aged ≥ 12 years | | | |
| **93** Painful menstruation[1] | 0 | □ Not anymore □ Intermittent □ Persistent | 1 2 3 |
| **94** Changes in sexual function[1] | 0 | □ Not anymore □ Intermittent □ Persistent | 1 2 3 |
| **95** Further urogenital complaints: | | | |

**96** Further complaints: ______________________________________________

______________________________________________

Adapted from:

[1] WHO. Global COVID-19 Clinical Platform Case Report Form (CRF) for Post COVID condition (Post COVID-19 CRF). 2021 Feb. Retrieved on 17.11.2021 von https://cdn.who.int/media/docs/default-source/3rd-edl-submissions/who_crf_postcovid_feb9_2021.pdf?sfvrsn=76afd14_1&download=true.

[2] DGPI. DGPI Survey – Long COVID-19. Retrieved on: 26.07.2021 von https://dgpi.de/post-covid-19-survey/.

[3] Sletten DM, Suarez GA, Low PA, Mandrekar J, Singer W. COMPASS 31: a refined and abbreviated Composite Autonomic Symptom Score. Mayo Clin Proc. 2012 Dec; 87(12):1196-201. doi: 10.1016/j.mayocp.2012.10.013.
